# A Tale of Two Models: Linear Regression and Poisson GLM Yield Identical Trends for COVID-19 Mortality in Italy

**DOI:** 10.1101/2025.09.11.25335570

**Authors:** Marco Roccetti, Giuseppe Cacciapuoti

## Abstract

While it is undisputed that Poisson GLM models represent the gold standard for counting COVID-19 deaths, recent studies have analyzed the seasonal growth and decline trends of these deaths in Italy using a simple segmented linear regression. They found that, despite an overall decreasing trend throughout the entire period analyzed (2021-2025), rising mortality trends from COVID-19 emerged in all summers and winters of the period, though they were more pronounced in winter. The technical reasons for the general unsuitability of using linear regression for the precise counting of deaths are well-known. Nevertheless, the question remains whether, under certain circumstances, the use of linear regression can provide a valid and useful tool in a specific context, for example, to highlight the slopes of seasonal growth/decline in deaths more quickly and clearly. Given this background, this paper presents a comparison between the use of linear regression and a Poisson GLM model with the aforementioned death data, leading to the following conclusions. Appropriate statistical hypothesis testing procedures have demonstrated that the conditions of a normal distribution of residuals, their homoscedasticity, and the lack of autocorrelation were essentially guaranteed in this particular Italian case (weekly COVID-19 deaths in Italy, from 2021 to 2025) with very rare exceptions, thus ensuring an acceptable performance of linear regression. This was further confirmed by the development of a Poisson GLM model, which showed: a) a 100% correspondence with the linear regression model in identifying the growth or decline trend across all 11 seasonal periods considered; b) that both the simple linear and Poisson models demonstrated a comparable great accuracy in counting COVID-19 deaths, with mean MAE values, per period, respectively of 88.60 and 62.76. Based on an average of approximately 6,630 deaths per period, the Poisson model showed a percentage error of 1.15%, while the linear regression model’s error was just 1.48%.

## 1. Introduction

In the field of statistical modeling, the fundamental premise of a rigorous analysis is the selection of a regression technique that aligns with the inherent characteristics of the data [1]. This is particularly critical when dealing with count data, which represent discrete, non-negative numbers, such as the number of events occurring over a fixed period. Unlike continuous variables that can take any value, count data are governed by distinct statistical properties. The widely adopted linear regression model, which operates under the assumption that the response variable is normally distributed, is fundamentally ill-suited for this type of data. The use of a linear model on count data can lead to serious statistical violations, including heteroscedasticity (non-constant variance) and the potential for predicting nonsensical, negative values, which are anathema to the very nature of counting [2].

For these reasons, the statistical community has long embraced Generalized Linear Models (GLMs), with Poisson regression standing as the undisputed gold standard for analyzing count data. Poisson GLMs are meticulously designed to accommodate the unique properties of discrete counts by employing a logarithmic link function that connects the linear predictor to the expected value of the counts [3, 4]. This ensures that all predictions remain non-negative which is a core reason for the model reliability. The Poisson model’s foundational assumption of equal dispersion (where the mean and variance are equal) makes it the most theoretically sound and widely accepted approach for count-based analysis in fields ranging from epidemiology to economics [5-7].

Despite this established statistical orthodoxy, recent research has explored an unconventional, yet compelling, application of a simpler technique. A recent study broke from convention by applying a segmented linear regression to analyze weekly COVID-19 death counts in Italy [8]. While statistically unorthodox, this methodology yielded powerful and highly interpretable results. The study, which investigated nine distinct seasonal periods (three winters, three summers, and three intermediate periods between winter and subsequent summer), revealed a clear and consistent pattern: all summers and winters within the analyzed period showed rising mortality trends, with the winter surges being consistently more pronounced, while the intermediate periods exhibited strong downward trends.

The true strength of this segmented linear approach laid in its ability to provide a visually impactful and intuitive representation of the mortality dynamics. By fitting straight lines to each seasonal segment, the model offered a clear and easily understandable visualization of the growth and decline rates through the slopes of the lines. This visual simplicity is a significant advantage, as it is often more accessible to a broader audience than the abstract logarithmic or exponential curves inherent in Poisson models.

The compelling findings of the initial research prompted a subsequent, confirmatory study to validate the discovered pattern over a (partially) new, extended period [9]. This work specifically extended the analysis to an entire, continuous year from May 2024 to May 2025 and was specifically designed to verify if the observed seasonal rhythm of growth during summer and winter, followed by a decline in the period from late winter to the end of the subsequent spring, would persist over a completely new 12-month span. This additional analysis, which built upon the techniques developed to analyze the original nine seasonal segments of [8], provided a more robust and comprehensive view of the phenomenon, demonstrating the durability of the observed COVID-19 mortality cycle and solidifying the linear model potential as a reliable tool for trend identification.

Given this background, a critical question comes now to the fore in all its significance: can the use of linear regression, despite its theoretical limitations, be considered an acceptable and even valuable tool in the unique context of seasonal COVID-19 mortality analysis, where its primary goal is to provide a quick and visually impactful representation of seasonal trends? This present paper aims to address this question by presenting a detailed, head-to-head comparison between a linear regression model and a Poisson GLM, using the same comprehensive dataset of Italian COVID-19 deaths from September 2021 to May 2025. The core of our investigation is not to dismiss the statistical rigor of GLMs, but rather to assess whether, under specific empirical conditions, a simpler model can provide comparable predictive power while offering superior interpretability and visual clarity. Our methodology involved two key steps. First, we conducted a series of rigorous statistical hypothesis tests on the linear regression model of [8, 9] to ensure its underlying assumptions were met. These tests assessed for the normal distribution of residuals, homoscedasticity (constant variance), and the absence of autocorrelation. Our findings reveal that, with very rare exceptions, these conditions were largely satisfied in this particular dataset, suggesting that the use of linear regression was indeed statistically tenable. Second, we developed a Poisson GLM on the same data to directly compare its performance against the linear model.

The results were remarkable: a) the Poisson model showed a 100% correspondence with the linear regression model in identifying the growth or decline trend across all eleven sub-periods (the nine from the original study plus the two additional seasons from the 2024/2025 period). Furthermore, both models demonstrated a comparable high level of accuracy in predicting death counts. The mean absolute error (MAE) for the linear model was 88.60, while the Poisson model MAE was 62.76. Based on an average of approximately 6,300 deaths per period, this translates to a relative error of only 1.48% for the linear model and 1.15% for the Poisson model.

These results have suggested that, in the context of analyzing Italian COVID-19 mortality data over this long period, the two models produced nearly identical outcomes. This finding highlights the practical utility of linear regression for this specific application, particularly for its ability to provide a clear and intuitive visual representation of seasonal trends through the slopes of its fitted lines, which are arguably more effective for communicating the phenomenon than the logarithmic/exponential curves inherent in Poisson models.

The remainder of this paper details the statistical procedures, models, and comparisons that led to these conclusions. In particular, in the next Section 2, we describe the source of the data used in this study and provide a comprehensive summary of all that data presented in tabular format. Furthermore, in the same Section, we provide the foundational information needed to discuss linear and Poisson regression, as well as the necessary background to understand the scope of the statistical tests conducted to verify the normality, homoscedasticity, and non-autocorrelation of residuals. In Section 3, we present the results obtained, both from the statistical tests and from the Poisson regression. Section 4, discusses a comparison between the outcomes of the linear and Poisson models, while Section 5 provides the conclusion of the work.

## 2. Materials and Methods

In this Section, we provide all the necessary details on the data and methods used in this study, allowing readers to easily replicate our results. We focused on the time series of Italian COVID-19 death data, which were subjected to various transformations, as described below.

### 2.1. Sources of COVID-19 Deaths Data

For the study of weekly COVID-19 deaths in Italy, all data were sourced from the two certified and currently available repositories within the country. These official sources are: i) the repository maintained by the Italian Civil Protection Department under the Italian Presidency of the Council of Ministers (https://github.com/pcm-dpc/COVID-19/blob/master/dati-andamento-nazionale), and ii) the repository maintained by the Italian Ministry of Health (https://www.salute.gov.it/new/it/tema/covid-19/report-set-timanali-covid-19/). These same data were used in our previous studies [8, 9] and cover a total of weeks of weekly COVID-19 deaths, spanning from September 23, 2021, to May 21, 2025. While the aforementioned are the primary sources, to make this study independently comprehensible, the time series of these weekly deaths, along with other relevant information, is also presented in a tabular format in the following Sections.

### 2.2. Linear Regression Fit to COVID-19 Deaths Data

We here summarize the results of fitting a linear regression model to the 210 weeks of COVID-19 death data from September 2021 to May 2025 mentioned above. The detailed findings were previously described in papers [8, 9], nonetheless the general principle behind this fitting procedure was to follow the trends of growth and decline in the historical time series of weekly deaths by dividing the entire period into seasonal segments. For each segment, the linear regression model identified the corresponding growth or decline trend, which is best expressed by the slope of the corresponding segment. This slope is captured by the well-known parameter *β_1_* in the classic linear regression equation [10]:

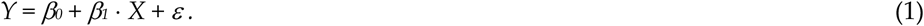

It is important to note that the definition of these seasonal segments was not strictly based on astronomical seasons. The segments were slightly adjusted, either lengthened or shortened, to better fit the regression lines, with the goal of keeping the goodness-of-fit, measured by the R^2^ parameter, around a 70% threshold on average. This approach allowed us to more accurately model the specific growth and decline phases of the pandemic throughout the analyzed period, yielding a total of nine seasonal segments.

To briefly summarize the main findings obtained by applying this procedure in [8], it should be recalled that the linear regression model has demonstrated that, throughout the examined three-year period (late 2021 to late 2024) dominated by the Omicron and post-Omicron variants: i) the overall trend of weekly COVID-19 mortality was in decline, yet ii) there were notable increases in deaths during all winters and summers. These rising mortality variations were more pronounced in winter than in summer. Conversely, deaths were less frequent in the intermediate periods between winter and summer. This study, therefore, concluded that essentially, although the general downward trend of COVID-19 mortality in Italy was favorable, transient rises in mortality occurred in both winter and summer but were largely offset by the consistent downward drifts during the intermediate seasons.

To further confirm these, in some respects, surprising results, particularly the ascending mortality trends in summer in addition to winter, a subsequent study was conducted to examine an entire year, from May 2024 to May 2025, using the same segmented linear regression model of [8]. This later study [9] also confirmed the ascending mortality trend, which began in summer, or late spring, and connected with the upward trend of autumn and the following winter. The trend then started to decline substantially from mid-winter all the way through spring. This descending period was precisely what had been identified as the *intermediate* period in the previously mentioned work [8]. Specifically, these two additional seasonal segments began and ended on dates May 16, 2024 and November 14, 2024, and November 21, 2024 and May 21, 2025, respectively, for a total duration of 53 weeks, thus constituting two new seasonal segments for study to be added to the previous 9, for a total of 11.

Table 1 summarizes these 11 periods, showing their duration in weeks, start and end dates, and season type.

**Table 1.** The 11 segments identified in Italy in the period from September 2021 to May 2025 with their seasonal connotation provided by the linear regression model of [8, 9].

| Seasonal Segment | Season Type | Number of weeks | Segment beginning | Segment end |
| --- | --- | --- | --- | --- |
| Winter 2021 | Fall-Winter | 19 | 9/23/21 | 1/28/22 |
| Intermediate 2022 | Extended Spring | 19 | 2/4/22 | 6/10/22 |
| Summer 2022 | Summer | 10 | 6/17/22 | 8/19/22 |
| Winter 2022 | Fall-Winter | 18 | 8/26/22 | 12/23/22 |
| Intermediate 2023 | Extended Spring | 27 | 12/30/22 | 6/30/23 |
| Summer 2023 | Summer | 10 | 7/7/23 | 9/7/23 |
| Winter 2023 | Fall-Winter | 16 | 9/14/23 | 12/28/23 |
| Intermediate 2024 | Extended Spring | 21 | 1/4/24 | 5/23/24 |
| Summer 2024 | Summer | 17 | 5/30/24 | 11/19/24 |
| Summer-Winter 2024-25 | Summer + Winter | 21 | 1/4/24 | 5/23/24 |
| Extended Spring 2025 | Extended Spring | 17 | 5/30/24 | 11/19/24 |

The following four Tables (2-5) provide a concise yet comprehensive overview of the data and results from the two previous studies [8, 9] over more than 210 weeks. The extensive time frame, spanning from September 2021 to May 2025, has been divided into four consecutive macro-periods (macro-periods 1, 2 3 and 4). Each macro-period 1-3 encompasses an autumn-winter phase (Winter), an intermediate phase (Intermediate), and the subsequent summer (Summer). For each macro-period, we presents in the second column of each Table the raw weekly COVID-19 death data. The subsequent columns of each Table provide the corresponding regression results, including the type of season, the ascending or descending trend identified by the regression, along with the *β_1_* coefficient, the point-estimate prediction (in terms of weekly deaths) provided by the linear regression model, and the residual, that is the difference between the actual and estimated death values. The residual’s sign is positive for overestimation and negative for underestimation. In summary, macro-period 1 (Table 2): from September 23, 2021, to August 18, 2022, 48 weeks; macro-period 2 (Table 3): from August 26, 2022, to September 7, 2023, 55 weeks; macro-period 3 (Table 4): from September 14, 2023, to September 19, 2024, 54 weeks.

**Table 2.**
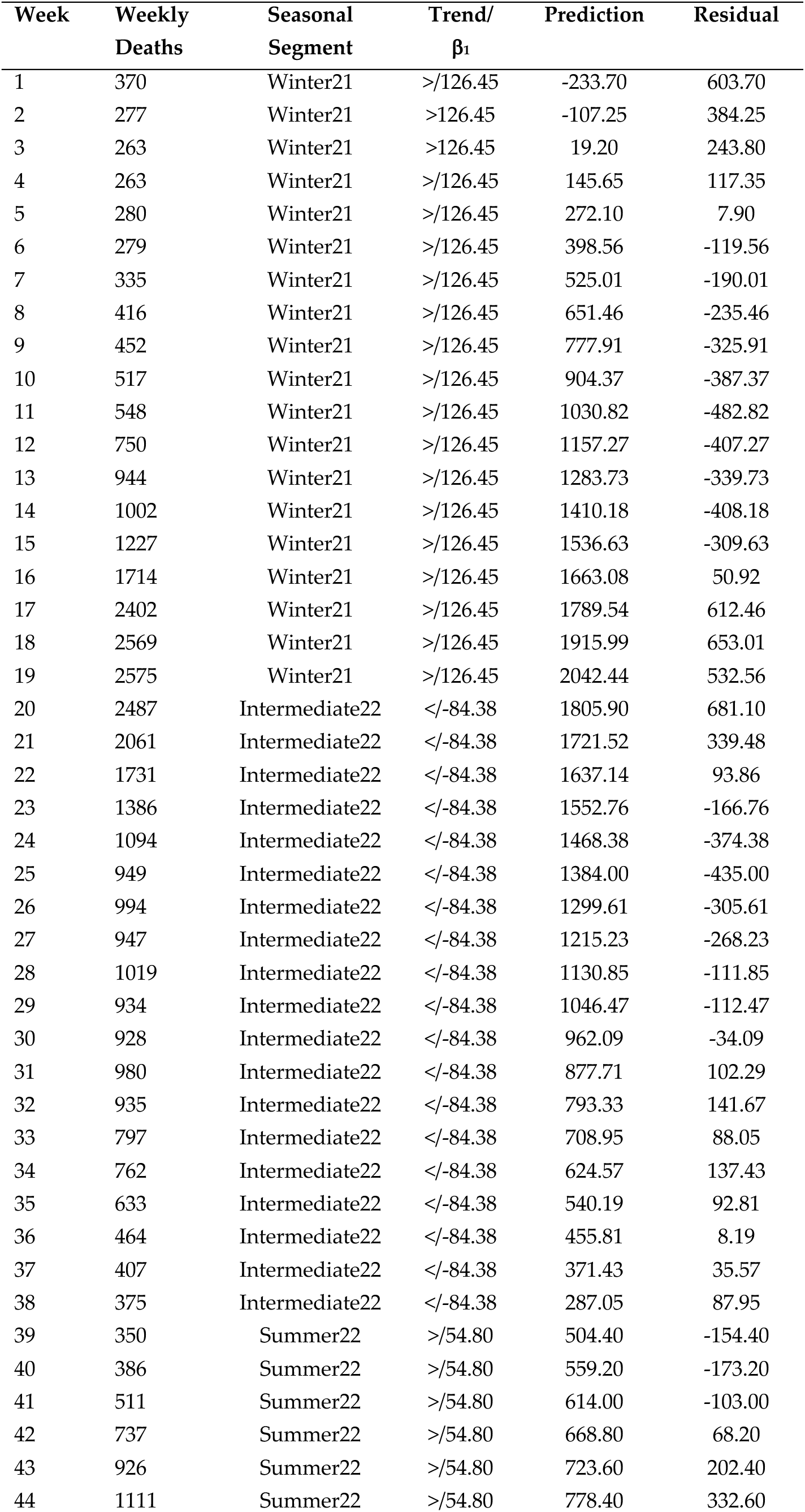
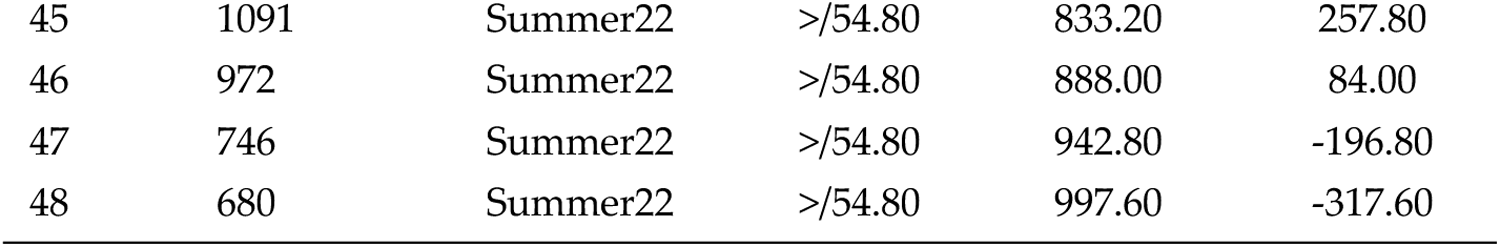
Macro-period 1 (September 23, 2021, to August 18, 2022). 48 consecutive weeks of COVID-19 deaths in Italy plus segmented linear regression results, including: Season type, Identified seasonal trend of growth (>) or decline (<) and segment slope *β_1_*, Predictions, Residuals.

| Week | Weekly Deaths | Seasonal Segment | Trend/<br>$\beta_1$ | Prediction | Residual |
| --- | --- | --- | --- | --- | --- |
| 1 | 370 | Winter21 | >/126.45 | -233.70 | 603.70 |
| 2 | 277 | Winter21 | >126.45 | -107.25 | 384.25 |
| 3 | 263 | Winter21 | >126.45 | 19.20 | 243.80 |
| 4 | 263 | Winter21 | >/126.45 | 145.65 | 117.35 |
| 5 | 280 | Winter21 | >/126.45 | 272.10 | 7.90 |
| 6 | 279 | Winter21 | >/126.45 | 398.56 | -119.56 |
| 7 | 335 | Winter21 | >/126.45 | 525.01 | -190.01 |
| 8 | 416 | Winter21 | >/126.45 | 651.46 | -235.46 |
| 9 | 452 | Winter21 | >/126.45 | 777.91 | -325.91 |
| 10 | 517 | Winter21 | >/126.45 | 904.37 | -387.37 |
| 11 | 548 | Winter21 | >/126.45 | 1030.82 | -482.82 |
| 12 | 750 | Winter21 | >/126.45 | 1157.27 | -407.27 |
| 13 | 944 | Winter21 | >/126.45 | 1283.73 | -339.73 |
| 14 | 1002 | Winter21 | >/126.45 | 1410.18 | -408.18 |
| 15 | 1227 | Winter21 | >/126.45 | 1536.63 | -309.63 |
| 16 | 1714 | Winter21 | >/126.45 | 1663.08 | 50.92 |
| 17 | 2402 | Winter21 | >/126.45 | 1789.54 | 612.46 |
| 18 | 2569 | Winter21 | >/126.45 | 1915.99 | 653.01 |
| 19 | 2575 | Winter21 | >/126.45 | 2042.44 | 532.56 |
| 20 | 2487 | Intermediate22 | </-84.38 | 1805.90 | 681.10 |
| 21 | 2061 | Intermediate22 | </-84.38 | 1721.52 | 339.48 |
| 22 | 1731 | Intermediate22 | </-84.38 | 1637.14 | 93.86 |
| 23 | 1386 | Intermediate22 | </-84.38 | 1552.76 | -166.76 |
| 24 | 1094 | Intermediate22 | </-84.38 | 1468.38 | -374.38 |
| 25 | 949 | Intermediate22 | </-84.38 | 1384.00 | -435.00 |
| 26 | 994 | Intermediate22 | </-84.38 | 1299.61 | -305.61 |
| 27 | 947 | Intermediate22 | </-84.38 | 1215.23 | -268.23 |
| 28 | 1019 | Intermediate22 | </-84.38 | 1130.85 | -111.85 |
| 29 | 934 | Intermediate22 | </-84.38 | 1046.47 | -112.47 |
| 30 | 928 | Intermediate22 | </-84.38 | 962.09 | -34.09 |
| 31 | 980 | Intermediate22 | </-84.38 | 877.71 | 102.29 |
| 32 | 935 | Intermediate22 | </-84.38 | 793.33 | 141.67 |
| 33 | 797 | Intermediate22 | </-84.38 | 708.95 | 88.05 |
| 34 | 762 | Intermediate22 | </-84.38 | 624.57 | 137.43 |
| 35 | 633 | Intermediate22 | </-84.38 | 540.19 | 92.81 |
| 36 | 464 | Intermediate22 | </-84.38 | 455.81 | 8.19 |
| 37 | 407 | Intermediate22 | </-84.38 | 371.43 | 35.57 |
| 38 | 375 | Intermediate22 | </-84.38 | 287.05 | 87.95 |
| 39 | 350 | Summer22 | >/54.80 | 504.40 | -154.40 |
| 40 | 386 | Summer22 | >/54.80 | 559.20 | -173.20 |
| 41 | 511 | Summer22 | >/54.80 | 614.00 | -103.00 |
| 42 | 737 | Summer22 | >/54.80 | 668.80 | 68.20 |
| 43 | 926 | Summer22 | >/54.80 | 723.60 | 202.40 |
| 44 | 1111 | Summer22 | >/54.80 | 778.40 | 332.60 |
| 45 | 1091 | Summer22 | >/54.80 | 833.20 | 257.80 |
| 46 | 972 | Summer22 | >/54.80 | 888.00 | 84.00 |
| 47 | 746 | Summer22 | >/54.80 | 942.80 | -196.80 |
| 48 | 680 | Summer22 | >/54.80 | 997.60 | -317.60 |

**Table 3.** Macro-period 2 (August 26, 2022, to September 7, 2023). 55 consecutive weeks of COVID-19 deaths in Italy plus segmented linear regression results, including: Season type, Identified seasonal trend of growth (>) or decline (<) and segment slope *β_1_*, Predictions, Residuals.

| Week | Weekly Deaths | Seasonal Segment | Trend/<br>$\beta_i$ | Prediction | Residual |
| --- | --- | --- | --- | --- | --- |
| 49 | 536 | Winter22 | >/23.28 | 330.72 | 205.28 |
| 50 | 435 | Winter22 | >/23.28 | 354.00 | 81.00 |
| 51 | 366 | Winter22 | >/23.28 | 377.29 | -11.29 |
| 52 | 311 | Winter22 | >/23.28 | 400.57 | -89.57 |
| 53 | 279 | Winter22 | >/23.28 | 423.85 | -144.85 |
| 54 | 302 | Winter22 | >/23.28 | 447.13 | -145.13 |
| 55 | 429 | Winter22 | >/23.28 | 470.41 | -41.41 |
| 56 | 574 | Winter22 | >/23.28 | 493.69 | 80.31 |
| 57 | 581 | Winter22 | >/23.28 | 516.97 | 64.03 |
| 58 | 496 | Winter22 | >/23.28 | 540.25 | -44.25 |
| 59 | 549 | Winter22 | >/23.28 | 563.53 | -14.53 |
| 60 | 533 | Winter22 | >/23.28 | 586.81 | -53.81 |
| 61 | 580 | Winter22 | >/23.28 | 610.09 | -30.09 |
| 62 | 635 | Winter22 | >/23.28 | 633.37 | 1.63 |
| 63 | 686 | Winter22 | >/23.28 | 656.65 | 29.35 |
| 64 | 719 | Winter22 | >/23.28 | 679.94 | 39.06 |
| 65 | 798 | Winter22 | >/23.28 | 703.21 | 94.79 |
| 66 | 706 | Winter22 | >/23.28 | 725.50 | -20.50 |
| 67 | 775 | Intermediate23 | </-17.76 | 462.91 | 312.09 |
| 68 | 576 | Intermediate23 | </-17.76 | 445.15 | 130.85 |
| 69 | 495 | Intermediate23 | </-17.76 | 427.39 | 67.61 |
| 70 | 345 | Intermediate23 | </-17.76 | 409.62 | -64.62 |
| 71 | 439 | Intermediate23 | </-17.76 | 391.86 | 47.14 |
| 72 | 279 | Intermediate23 | </-17.76 | 374.10 | -95.10 |
| 73 | 299 | Intermediate23 | </-17.76 | 356.34 | -57.34 |
| 74 | 244 | Intermediate23 | </-17.76 | 338.57 | -94.57 |
| 75 | 228 | Intermediate23 | </-17.76 | 320.81 | -92.81 |
| 76 | 216 | Intermediate23 | </-17.76 | 330.72 | -87.805 |
| 77 | 212 | Intermediate23 | </-17.76 | 285.29 | -73.29 |
| 78 | 183 | Intermediate23 | </-17.76 | 267.52 | -84.52 |
| 79 | 156 | Intermediate23 | </-17.76 | 249.76 | -93.76 |
| 80 | 173 | Intermediate23 | </-17.76 | 232.00 | -59.00 |
| 81 | 129 | Intermediate23 | </-17.76 | 214.24 | -85.24 |
| 82 | 191 | Intermediate23 | </-17.76 | 196.47 | -5.47 |
| 83 | 156 | Intermediate23 | </-17.76 | 178.71 | -22.71 |
| 84 | 166 | Intermediate23 | </-17.76 | 160.95 | 5.05 |
| 85 | 176 | Intermediate23 | </-17.76 | 143.19 | 32.81 |
| 86 | 162 | Intermediate23 | </-17.76 | 125.42 | 36.58 |
| 87 | 150 | Intermediate23 | </-17.76 | 107.66 | 42.34 |
| 88 | 125 | Intermediate23 | </-17.76 | 89.90 | 35.10 |
| 89 | 108 | Intermediate23 | </-17.76 | 72.14 | 35.86 |
| 90 | 81 | Intermediate23 | </-17.76 | 54.37 | 26.63 |
| 91 | 76 | Intermediate23 | </-17.76 | 36.61 | 39.39 |
| 92 | 86 | Intermediate23 | </-17.76 | 18.85 | 67.15 |
| 93 | 38 | Intermediate23 | </-17.76 | 1.09 | 36.91 |
| 94 | 36 | Summer23 | >/6.72 | 26.73 | 9.27 |
| 95 | 45 | Summer23 | >/6.72 | 33.45 | 11.55 |
| 96 | 25 | Summer23 | >/6.72 | 40.18 | -15.18 |
| 97 | 41 | Summer23 | >/6.72 | 46.91 | -5.91 |
| 98 | 65 | Summer23 | >/6.72 | 53.64 | 11.48 |
| 99 | 56 | Summer23 | >/6.72 | 60.36 | -4.36 |
| 100 | 44 | Summer23 | >/6.72 | 67.09 | -23.09 |
| 101 | 65 | Summer23 | >/6.72 | 73.82 | -8.82 |
| 102 | 94 | Summer23 | >/6.72 | 80.54 | 13.46 |
| 103 | 99 | Summer23 | >/6.72 | 87.27 | 11.73 |

**Table 4.**
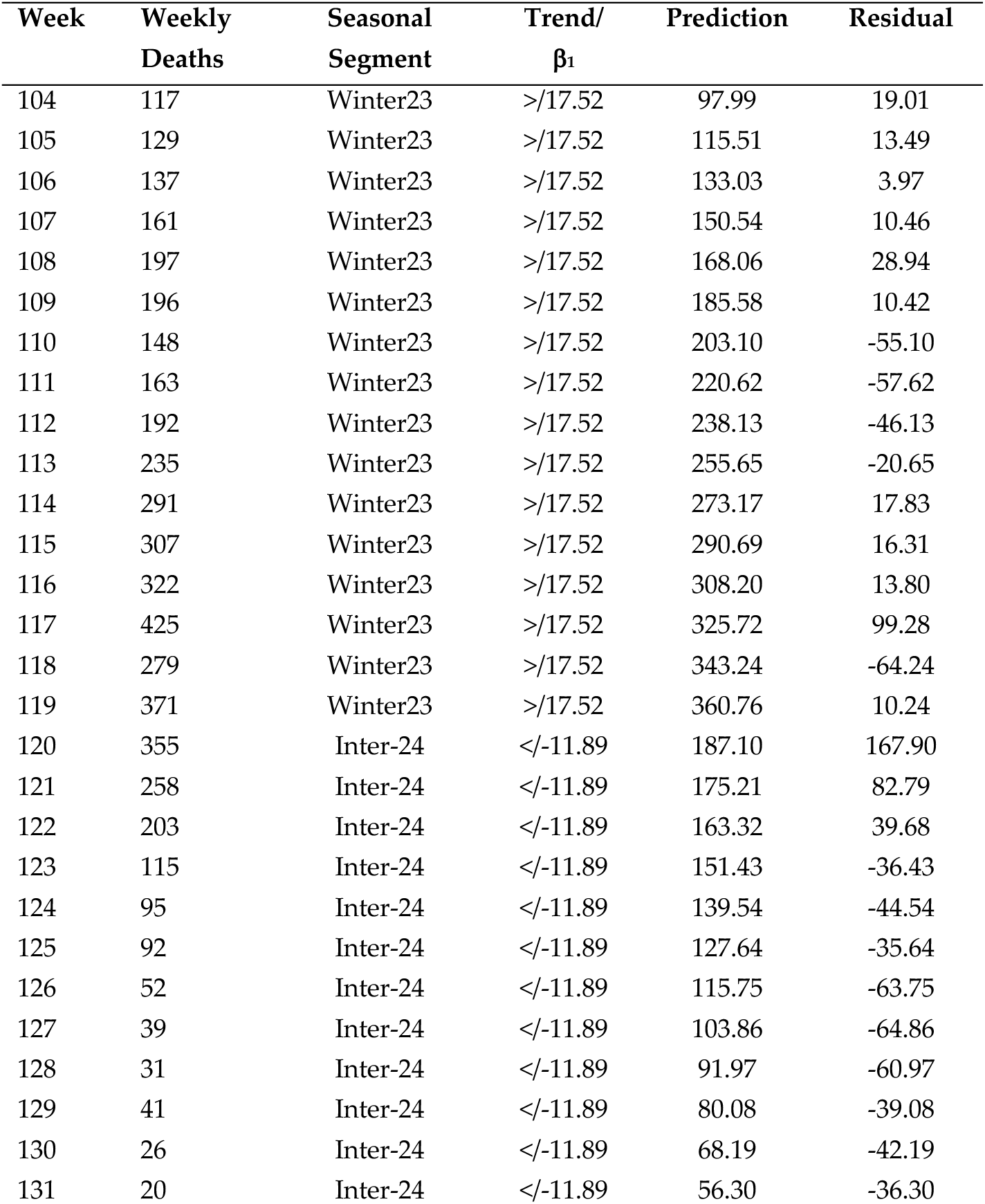
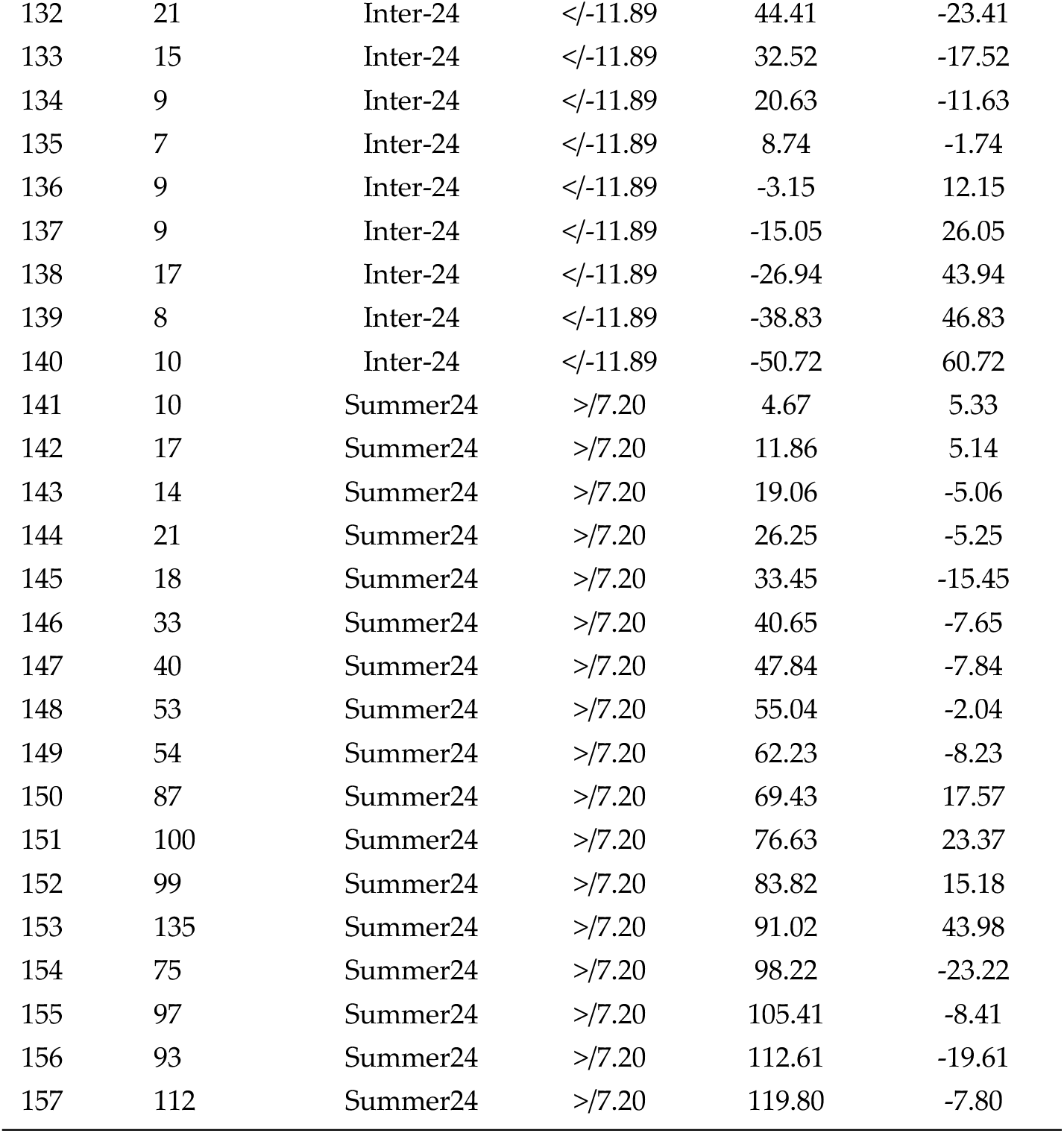
Macro-period 3 (from September 14, 2023, to September 19, 2024). 54 consecutive weeks of COVID-19 deaths in Italy plus segmented linear regression results, including: Season type, Identified seasonal trend of growth (>) or decline (<) and segment slope *β_1_*, Predictions, Residuals.

| Week | Weekly Deaths | Seasonal Segment | Trend/<br>$\beta_1$ | Prediction | Residual |
| --- | --- | --- | --- | --- | --- |
| 104 | 117 | Winter23 | >/17.52 | 97.99 | 19.01 |
| 105 | 129 | Winter23 | >/17.52 | 115.51 | 13.49 |
| 106 | 137 | Winter23 | >/17.52 | 133.03 | 3.97 |
| 107 | 161 | Winter23 | >/17.52 | 150.54 | 10.46 |
| 108 | 197 | Winter23 | >/17.52 | 168.06 | 28.94 |
| 109 | 196 | Winter23 | >/17.52 | 185.58 | 10.42 |
| 110 | 148 | Winter23 | >/17.52 | 203.10 | -55.10 |
| 111 | 163 | Winter23 | >/17.52 | 220.62 | -57.62 |
| 112 | 192 | Winter23 | >/17.52 | 238.13 | -46.13 |
| 113 | 235 | Winter23 | >/17.52 | 255.65 | -20.65 |
| 114 | 291 | Winter23 | >/17.52 | 273.17 | 17.83 |
| 115 | 307 | Winter23 | >/17.52 | 290.69 | 16.31 |
| 116 | 322 | Winter23 | >/17.52 | 308.20 | 13.80 |
| 117 | 425 | Winter23 | >/17.52 | 325.72 | 99.28 |
| 118 | 279 | Winter23 | >/17.52 | 343.24 | -64.24 |
| 119 | 371 | Winter23 | >/17.52 | 360.76 | 10.24 |
| 120 | 355 | Inter-24 | </-11.89 | 187.10 | 167.90 |
| 121 | 258 | Inter-24 | </-11.89 | 175.21 | 82.79 |
| 122 | 203 | Inter-24 | </-11.89 | 163.32 | 39.68 |
| 123 | 115 | Inter-24 | </-11.89 | 151.43 | -36.43 |
| 124 | 95 | Inter-24 | </-11.89 | 139.54 | -44.54 |
| 125 | 92 | Inter-24 | </-11.89 | 127.64 | -35.64 |
| 126 | 52 | Inter-24 | </-11.89 | 115.75 | -63.75 |
| 127 | 39 | Inter-24 | </-11.89 | 103.86 | -64.86 |
| 128 | 31 | Inter-24 | </-11.89 | 91.97 | -60.97 |
| 129 | 41 | Inter-24 | </-11.89 | 80.08 | -39.08 |
| 130 | 26 | Inter-24 | </-11.89 | 68.19 | -42.19 |
| 131 | 20 | Inter-24 | </-11.89 | 56.30 | -36.30 |
| 132 | 21 | Inter-24 | </-11.89 | 44.41 | -23.41 |
| 133 | 15 | Inter-24 | </-11.89 | 32.52 | -17.52 |
| 134 | 9 | Inter-24 | </-11.89 | 20.63 | -11.63 |
| 135 | 7 | Inter-24 | </-11.89 | 8.74 | -1.74 |
| 136 | 9 | Inter-24 | </-11.89 | -3.15 | 12.15 |
| 137 | 9 | Inter-24 | </-11.89 | -15.05 | 26.05 |
| 138 | 17 | Inter-24 | </-11.89 | -26.94 | 43.94 |
| 139 | 8 | Inter-24 | </-11.89 | -38.83 | 46.83 |
| 140 | 10 | Inter-24 | </-11.89 | -50.72 | 60.72 |
| 141 | 10 | Summer24 | >/7.20 | 4.67 | 5.33 |
| 142 | 17 | Summer24 | >/7.20 | 11.86 | 5.14 |
| 143 | 14 | Summer24 | >/7.20 | 19.06 | -5.06 |
| 144 | 21 | Summer24 | >/7.20 | 26.25 | -5.25 |
| 145 | 18 | Summer24 | >/7.20 | 33.45 | -15.45 |
| 146 | 33 | Summer24 | >/7.20 | 40.65 | -7.65 |
| 147 | 40 | Summer24 | >/7.20 | 47.84 | -7.84 |
| 148 | 53 | Summer24 | >/7.20 | 55.04 | -2.04 |
| 149 | 54 | Summer24 | >/7.20 | 62.23 | -8.23 |
| 150 | 87 | Summer24 | >/7.20 | 69.43 | 17.57 |
| 151 | 100 | Summer24 | >/7.20 | 76.63 | 23.37 |
| 152 | 99 | Summer24 | >/7.20 | 83.82 | 15.18 |
| 153 | 135 | Summer24 | >/7.20 | 91.02 | 43.98 |
| 154 | 75 | Summer24 | >/7.20 | 98.22 | -23.22 |
| 155 | 97 | Summer24 | >/7.20 | 105.41 | -8.41 |
| 156 | 93 | Summer24 | >/7.20 | 112.61 | -19.61 |
| 157 | 112 | Summer24 | >/7.20 | 119.80 | -7.80 |

The final Table 5 details the one-year period from May 2024 to May 2025, as studied in [9]. This study, also conducted using our linear regression model, aimed to confirm the ascending summer-autumn-winter COVID-19 mortality trend and the subsequent post-winter decline over an annual timeframe. It should be noted that the first 35 weeks of this final macro-period (the fourth) overlap with the third macro-period. For this macro-period 4 as well, the second column of Table 5 reports the weekly COVID-19 deaths, while the subsequent columns of Table 5 presents the linear regression model results: the season type, the growth or decline trend of mortality, with the corresponding *β_1_* values, the prediction values, and the residuals. Hence, the macro-period 4 (Table 5), from May 16, 2024, to May 21, 2025, for a duration of 53 weeks.

**Table 5.** Macro-period 4 (from May 16, 2024 to May 21, 2025). A full year of COVID-19 deaths in Italy (53 weeks) plus segmented linear regression results, including: Season type, Identified seasonal trend of growth (>) or decline (<), segment slope *β_1_*, Predictions, Residuals.

| Week | Weekly Deaths | Seasonal Segment | Trend/<br>$\beta_1$ | Prediction | Residual |
| --- | --- | --- | --- | --- | --- |
| 1 | 8 | Summer-Winter | >/4.08 | 15.25 | -7.25 |
| 2 | 10 | Summer-Winter | >/4.08 | 19.33 | -9.33 |
| 3 | 10 | Summer-Winter | >/4.08 | 23.41 | -13.41 |
| 4 | 17 | Summer-Winter | >/4.08 | 27.50 | -10.50 |
| 5 | 14 | Summer-Winter | >/4.08 | 31.58 | -17.58 |
| 6 | 21 | Summer-Winter | >/4.08 | 35.66 | -14.66 |
| 7 | 18 | Summer-Winter | >/4.08 | 39.75 | -21.75 |
| 8 | 33 | Summer-Winter | >/4.08 | 43.83 | -10.83 |
| 9 | 40 | Summer-Winter | >/4.08 | 47.91 | -7.91 |
| 10 | 53 | Summer-Winter | >/4.08 | 52.00 | 1.00 |
| 11 | 54 | Summer-Winter | >/4.08 | 56.08 | -2.08 |
| 12 | 87 | Summer-Winter | >/4.08 | 60.17 | 26.83 |
| 13 | 100 | Summer-Winter | >/4.08 | 64.25 | 35.75 |
| 14 | 99 | Summer-Winter | >/4.08 | 68.33 | 30.67 |
| 15 | 135 | Summer-Winter | >/4.08 | 72.42 | 62.58 |
| 16 | 75 | Summer-Winter | >/4.08 | 76.50 | -1.50 |
| 17 | 97 | Summer-Winter | >/4.08 | 80.58 | 16.42 |
| 18 | 93 | Summer-Winter | >/4.08 | 84.67 | 8.33 |
| 19 | 112 | Summer-Winter | >/4.08 | 88.75 | 23.25 |
| 20 | 85 | Summer-Winter | >/4.08 | 92.83 | -7.83 |
| 21 | 100 | Summer-Winter | >/4.08 | 96.92 | 3.08 |
| 22 | 117 | Summer-Winter | >/4.08 | 101.00 | 16.00 |
| 23 | 116 | Summer-Winter | >/4.08 | 105.09 | 10.91 |
| 24 | 108 | Summer-Winter | >/4.08 | 109.17 | -1.17 |
| 25 | 96 | Summer-Winter | >/4.08 | 113.25 | -17.25 |
| 26 | 86 | Summer-Winter | >/4.08 | 117.34 | -31.34 |
| 27 | 61 | Summer-Winter | >/4.08 | 121.42 | -60.42 |
| 28 | 47 | Extended Spring | </-1.81 | 48.49 | -1.49 |
| 29 | 46 | Extended Spring | </-1.81 | 46.67 | -0.67 |
| 30 | 44 | Extended Spring | </-1.81 | 44.86 | -0.86 |
| 31 | 43 | Extended Spring | </-1.81 | 43.04 | -0.04 |
| 32 | 29 | Extended Spring | </-1.81 | 41.23 | -12.23 |
| 33 | 31 | Extended Spring | </-1.81 | 39.41 | -8.41 |
| 34 | 45 | Extended Spring | </-1.81 | 37.60 | 7.40 |
| 35 | 44 | Extended Spring | </-1.81 | 35.79 | 8.21 |
| 36 | 58 | Extended Spring | </-1.81 | 33.97 | 24.03 |
| 37 | 43 | Extended Spring | </-1.81 | 32.16 | 10.84 |
| 38 | 24 | Extended Spring | </-1.81 | 30.34 | -6.34 |
| 39 | 25 | Extended Spring | </-1.81 | 28.53 | -3.53 |
| 40 | 29 | Extended Spring | </-1.81 | 26.71 | 2.29 |
| 41 | 13 | Extended Spring | </-1.81 | 24.90 | -11.90 |
| 42 | 17 | Extended Spring | </-1.81 | 23.09 | -6.09 |
| 43 | 25 | Extended Spring | </-1.81 | 21.27 | 3.73 |
| 44 | 16 | Extended Spring | </-1.81 | 19.46 | -3.46 |
| 45 | 17 | Extended Spring | </-1.81 | 17.64 | -0.64 |
| 46 | 20 | Extended Spring | </-1.81 | 15.83 | 4.17 |
| 47 | 6 | Extended Spring | </-1.81 | 14.01 | -8.01 |
| 48 | 8 | Extended Spring | </-1.81 | 12.20 | -4.20 |
| 49 | 9 | Extended Spring | </-1.81 | 10.39 | -1.39 |
| 50 | 1 | Extended Spring | </-1.81 | 8.57 | -7.57 |
| 51 | 13 | Extended Spring | </-1.81 | 6.76 | 6.24 |
| 52 | 13 | Extended Spring | </-1.81 | 4.94 | 8.06 |
| 53 | 5 | Extended Spring | </-1.81 | 3.13 | 1.87 |

In the end, not counting the 35-week overlap between macro-periods 3 and 4, the total number of weeks on which the linear regression model was fitted is 210. Of these, 157 were studied in [8] and 53 in [9].

### 2.3. Statistical Tests for Linear Regression on COVID-19 Mortality Data

With the linear regression model applied to the COVID-19 death data series reported in Tables 1-4, and the consequent values of the predictions and residuals, we can now perform a series of statistical tests. These tests, primarily applied to the residuals, that is the difference between predicted values and actually COVID-19 observed deaths, are essential for verifying whether the key conditions for a reliable applicability of a linear regression model have been met. Specifically, these tests check for the normality of the residual distribution, their homoscedasticity and the absence of autocorrelation. The rationale is that we use the following statistical tests to check if the residuals satisfy these conditions, allowing for a safe and valid use of the regression model used in [8, 9].

In particular, to test residuals for normality, we used both the Shapiro-Wilk test and the QQ plot procedure. The Shapiro-Wilk test is the principal method used to formally assess whether the residuals of a linear regression model are normally distributed [11]. The assumption of normality of residuals is crucial for the validity of hypothesis tests and the construction of confidence intervals for the linear model coefficients. This test calculates a statistic *W* which quantifies how well the sample data aligns with a normal distribution. The null hypothesis of the test is that the residuals are drawn from a normal distribution. A *p-value* from the test that is greater than a chosen significance level (we have chosen 0.05) indicates that the null hypothesis cannot be rejected, suggesting that the residuals are likely normally distributed.

To strengthen the verification of residual normality, a more empirical approach is usually employed, based on the visual inspection of a plot. We are talking about a Q-Q Plot [12]. A Quantile-Quantile (Q-Q) plot, in fact, provides a visual method for checking the normality of a model’s residuals, serving as an important complement to formal numerical statistical tests. The plot compares the ordered values of the residuals against the theoretical quantiles of a normal distribution. If the residuals are approximately normally distributed, the points on the plot will closely follow a straight diagonal line. Any significant deviation from this straight line or points that fall away from the line suggests that the residuals are not normally distributed, indicating a violation of the assumption. One common convention when using Q-Q plots is also to include a 95% confidence band around the diagonal line. The idea is that if the residuals are indeed normally distributed, the vast majority of the plot’s points should fall within this band.

A second fundamental condition for the reliable application of a linear regression model is the homoscedasticity of its residuals. This term refers to the property when the variance of the residuals is constant and uniform across all values of the independent variables. In other words, the spread of the data points around the regression line remains consistent throughout the entire range of predicted values. The violation of this condition, known as heteroscedasticity, renders the standard errors and significance tests unreliable, compromising the model’s conclusions. The Breusch-Pagan test is the specific statistical tool used to verify this condition, detecting whether the variance of the residuals is related to the independent variables [13]. In particular, the test works calculating an *LM* statistic which measures if the variance of a linear model’s residuals is constant. It is computed by running an auxiliary regression of the squared residuals on the original independent variables. The statistic is derived from this auxiliary regression’s R^2^ and follows a chi-squared distribution. A low p-value (typically < 0.05) for the *LM* statistic indicates heteroscedasticity, meaning the variance of the residuals is not constant which is exactly the situation we would want to avoid.

In the end, for a proper application of linear regression to counting problems, it is also essential to verify that the residuals are not autocorrelated. This serial correlation, where the error term of one observation is related to the next, is a common issue with time-series data, like weekly death counts. Its presence violates a core assumption of linear regression, leading to biased standard errors and unreliable hypothesis tests. This can cause to incorrectly conclude that a predictor is significant. Therefore, checking for auto-correlation is a crucial step to ensure the model’s findings are trustworthy. This verification can be done using ACF and PACF diagrams for autocorrelation. In simple terms, ACF (Autocorrelation Function) and PACF (Partial Autocorrelation Function) correlograms are powerful tools for detecting autocorrelation in the residuals of a regression model, particularly with time-series data [14]. Autocorrelation occurs when the residuals are not independent of each other. The ACF plot shows the correlation between a time series and its lagged values, helping to identify how long a residual’s effect persists. The PACF plot shows the direct correlation between a residual and its lagged values, after accounting for the influence of intermediate lags. Significant spikes on these plots (both ACF and PACF), extending beyond the confidence bands, are clear indicators of autocorrelation, suggesting the need for a different model or a transformation of the data.

### 2.4. Developing a Poisson GLM Model for COVID-19 Deaths Data

When analyzing weekly COVID-19 death counts, the Poisson Generalized Linear Model (GLM) is often a more suitable choice than linear regression. While linear regression assumes residuals are normally distributed, the Poisson GLM is specifically designed for discrete, non-negative count variables, assuming a Poisson distribution [15-19]. The model uses a link function (the natural logarithm) to transform the nonlinear relationship into a linear form. The model formula, applied to our case with a single time variable, is:

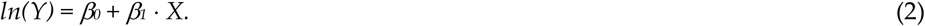

Here, *Y* represents the expected (or predicted) count of deaths, while *β_1_* is the regression coefficient. This approach produces an exponential curve that better reflects the natural growth and decline phases of COVID-19 deaths. To compare the two models (Poisson vs. Linear), it is then crucial to understand the meaning of *β_1_* in each. In linear regression, *β_1_* indicates a constant, additive change in the number of deaths for each new week. For example, a *β_1_* of *K* would mean an increase of *K* deaths every week. In the Poisson model, instead, *β_1_* represents a change in the logarithm of the expected count. To interpret this in terms of deaths, we need to calculate *e^β^*^1^. The result is a multiplicative factor: for instance, an *e^β^*^1^ of 1.05 indicates a 5% increase in deaths per week. This interpretation naturally aligns very well with the exponential growth and decay rates that characterize pandemics.

In closing, to determine which model provides a better seasonal profile, or whether they are ultimately comparable under the specific circumstances of our case, we have taken two approaches. In the first, we checked for any deviations in the seasonal predictions of COVID-19 mortality growth or decline, and their respective *β_1_* values (of course, after applying the link function in the case of Poisson). As a second check, we directly compared the predictions of the two models using the Mean Absolute Error (MAE). As is well known, MAE measures a model’s predictive accuracy by calculating the average of the absolute differences between predicted *Y* and observed *Ŷ* values with the formula:

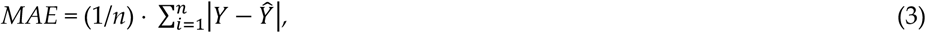

where *Y* is the actual number of weekly deaths and *Ŷ* is the predicted count. A lower MAE signifies a more accurate model. By calculating the MAE for both models for each season, we can see which model provides the better fit for the data during that specific seasonal period.

## 3. Results

This Results Section presents two types of achievements from the current study. First, the results related to the application of the various statistical tests mentioned previously to verify if the linear regression model developed in [8, 9] could be conducted with the assurance that the statistical assumptions of residual normality, their homoscedasticity, and the absence of autocorrelation were respected. Then, the second part of the results, shows the product of the Poisson regression, showing, for each of the 11 previously identified seasonal segments, the corresponding curves that identify the trend of COVID-19 mortality, along with the corresponding predictions in terms of COVID-19 deaths.

### 3.1. Results from the Statistical Validation of the Linear Regression Model for COVID-19 Mortality Data in Italy (2021-2025)

This part is divided into three types of results from corresponding checks that answer to the three subsequent questions: were the residuals of the linear model applied to the data of interest normally distributed across all 11 seasonal segments considered in the period 2021-2025? Were these residuals homoscedastic? Lastly, were they autocorrelated? First, we provide results of the statical test conducted for checking the residuals normality. The following Table 7 provides the results of the Shapiro-Wilk test on the 11 seasonal segments of interest, listing the *W* statistic (where it is recalled that the closer it is to 1, the more the hypothesis of residual normality holds), the p-value, and the outcome of the test.

**Table 7.**
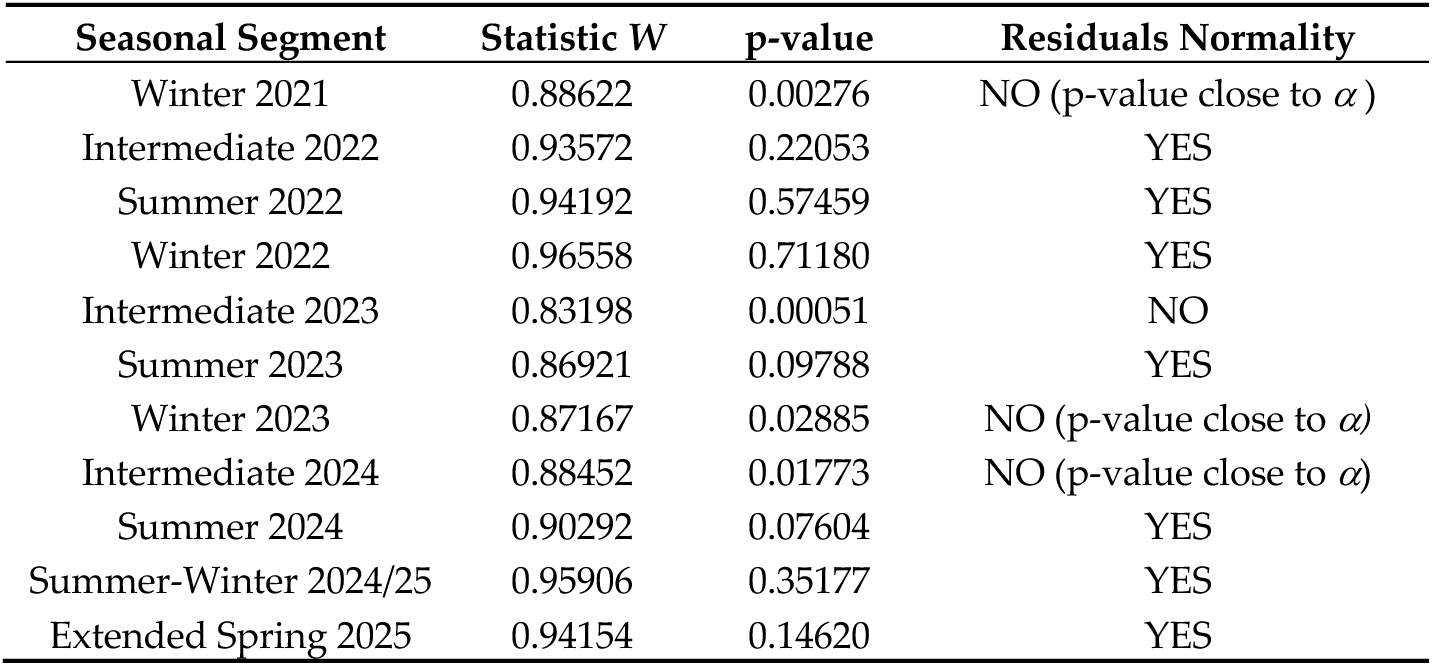
Shapiro-Wilk test results for the normality of residuals of the linear regression model for the seasonal segments in the period 2021 – 2025 (*α* = 0.05).

| Seasonal Segment | Statistic $W$ | p-value | Residuals Normality |
| --- | --- | --- | --- |
| Winter 2021 | 0.88622 | 0.00276 | NO (p-value close to $\alpha$) |
| Intermediate 2022 | 0.93572 | 0.22053 | YES |
| Summer 2022 | 0.94192 | 0.57459 | YES |
| Winter 2022 | 0.96558 | 0.71180 | YES |
| Intermediate 2023 | 0.83198 | 0.00051 | NO |
| Summer 2023 | 0.86921 | 0.09788 | YES |
| Winter 2023 | 0.87167 | 0.02885 | NO (p-value close to $\alpha$) |
| Intermediate 2024 | 0.88452 | 0.01773 | NO (p-value close to $\alpha$) |
| Summer 2024 | 0.90292 | 0.07604 | YES |
| Summer-Winter 2024/25 | 0.95906 | 0.35177 | YES |
| Extended Spring 2025 | 0.94154 | 0.14620 | YES |

What is noteworthy about these results is that either the test confirms the normality of the residuals (7 out of 11 cases), or the p-value, while below the significance threshold, is very close to it, always with *W* statistic values very near to 1. The four cases of failure of the normality test, due to their configuration, convinced us to plot the corresponding Q-Q plots for all 11 seasonal segments to get a better visual of this situation.

The following are therefore four Figures (Figures 1 to 4) that provide the Q-Q plots for all the winter segments of the investigated period (Figure 1), for all the summer segments (Figure 2), and for all the segments of the intermediate periods (Figure 3). Finally, Figure 4 provides the Q-Q plots for the two seasonal segments related to [9].

**Figure 1.**
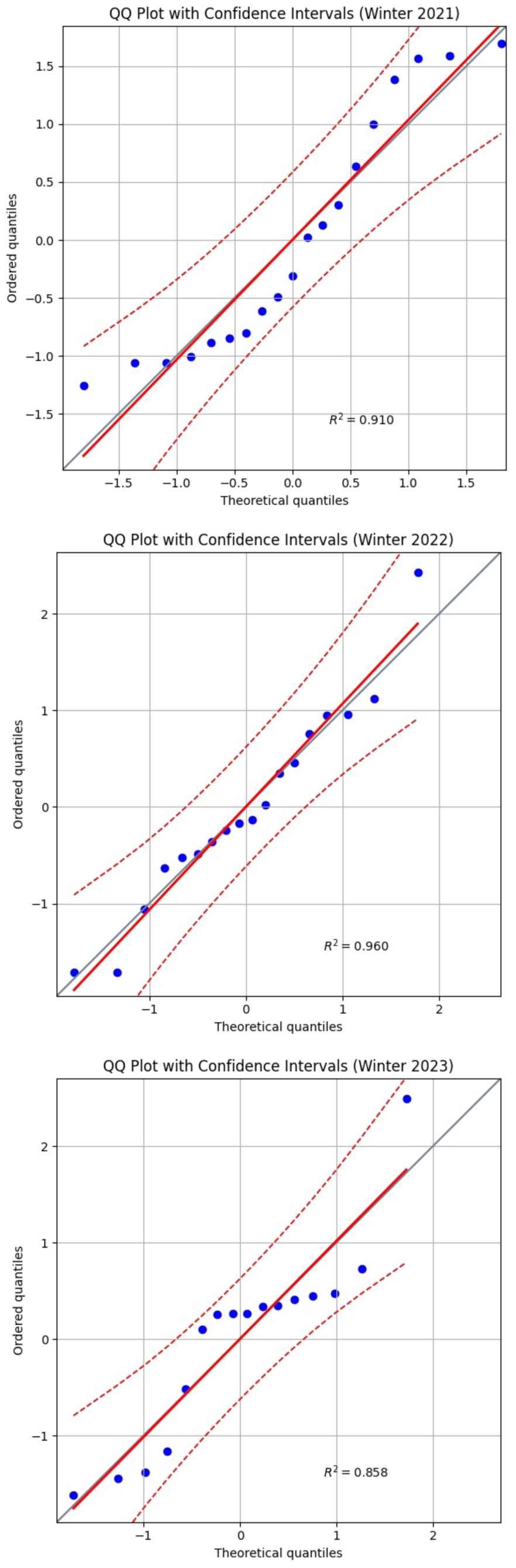
Q-Q Plots with a confidence intervals band of 95% for Winter 2021 (Top), Winter 2022 (Middle), Winter 2023 (Bottom).

**Figure 2.**
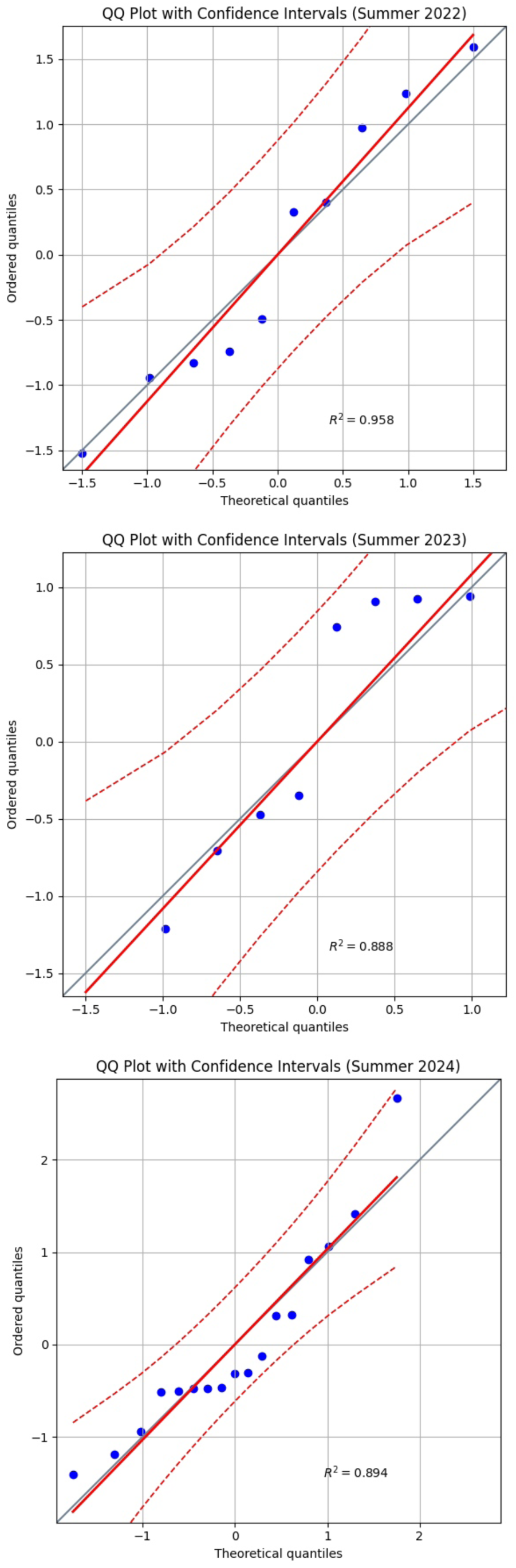
Q-Q Plots with a confidence intervals band of 95% for Summer 2022 (Top), Summer 2023 (Middle), Winter 2024 (Bottom).

**Figure 3.**
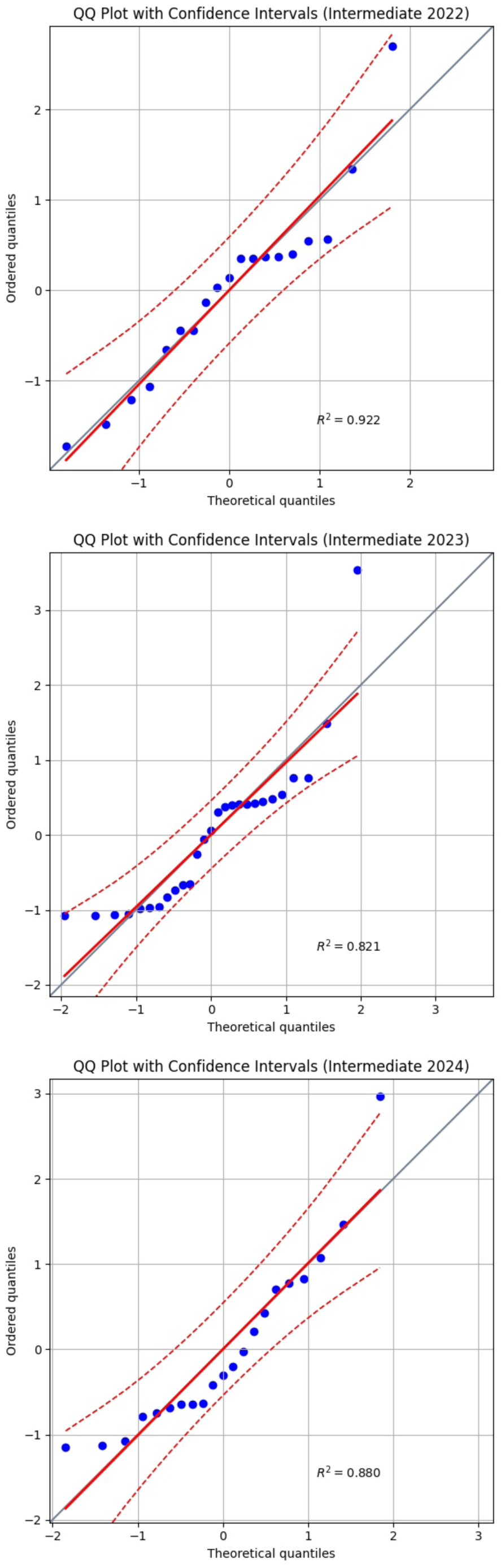
Q-Q Plots with a confidence intervals band of 95% for Intermediate 2022 (Top), Intermediate 2023 (Middle), Intermediate 2024 (Bottom).

**Figure 4.**
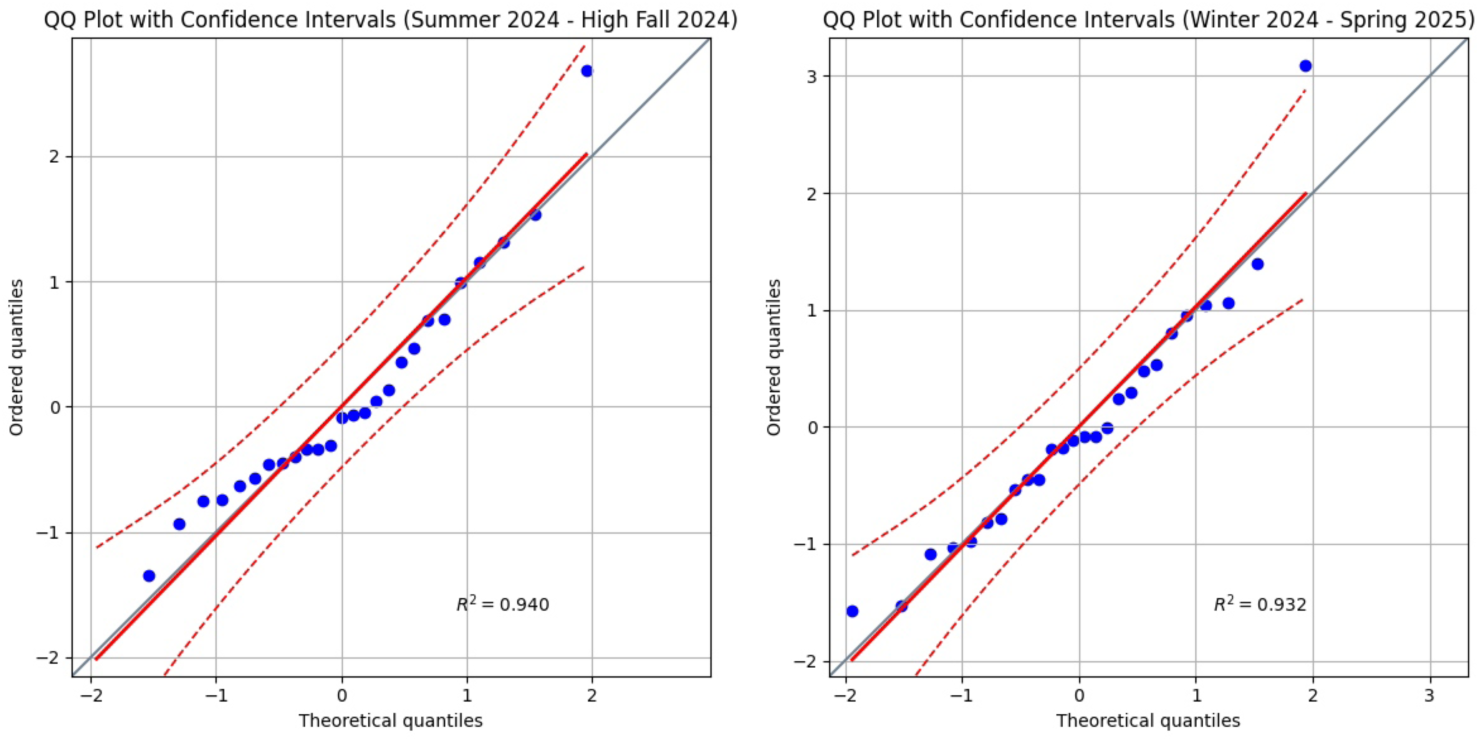
Q-Q Plots with a confidence intervals band of 95% for Summer-Winter 2024/25 (Left) and Extended Spring 2025 (Right).

As is easy to infer from a visual analysis of all the 11 Q-Q plots, the plot’s line aligns very well with the graph bisector. Most importantly, apart from three single points respectively in the plots for Winter 21, Intermediate 23 and Intermediate 24, the vast majority of points fall within the 95% confidence interval bands.

This demonstrates that, in addition to the decent normality results achieved with the more formal Shapiro-Wilk tests, from a practical numerical standpoint, all 11 seasonal segments exhibit a residual behavior that is almost entirely comparable to normality, with rare exceptions that deviate only slightly, and can therefore be defined as quasi-normal.

In conclusion, with 7 cases of normality and 4 very borderline, even if the entire spectrum of the 11 segments cannot be defined as perfectly normal, a situation largely close to normality can be deduced. Therefore, this certainly does not constitute one of the reasons for an unreliable application of the linear regression model.

Now it is time to check the homoscedasticity of the residuals produced by the linear regression model applied in [8, 9]. For this purpose, we show the results of the Breusch-Pagan tests for all 11 seasonal segments in the following Table 8, remembering that heteroscedasticity is the opposite of homoscedasticity.

**Table 8.** Breusch-Pagan test results for the heteroscedasticity of residuals of the seasonal segments in the period 2021 – 2025 (*α* = 0.05).

| Seasonal Segment | Statistics LM | p-value | Heteroscedasticity |
| --- | --- | --- | --- |
| Winter 2021 | 2.9880 | 0.08388 | No |
| Intermediate 2022 | 7.0823 | 0.00778 | Yes |
| Summer 2022 | 2.3209 | 0.12764 | No |
| Winter 2022 | 5.3495 | 0.02020 | No |
| Intermediate 2023 | 5.4031 | 0.02010 | No |
| Summer 2023 | 0.3138 | 0.57535 | No |
| Winter 2023 | 2.45566 | 0.11710 | No |
| Intermediate 2024 | 4.43587 | 0.03519 | Yes (p-value close to $\alpha$) |
| Summer 2024 | 2.42003 | 0.11979 | No |
| Summer-Winter 2024/25 | 2.12175 | 0.14522 | No |
| Extended Spring 2025 | 0.52298 | 0.46957 | No |

A quick look at this Table confirms that in 9 out of 11 cases, the residuals of the seasonal segments do not show heteroscedasticity, thus being homoscedastic.

For the Intermediate 2024 segment, an additional analysis of the residuals vs. fitted values plot (not reported here for the sake of conciseness) suggests the result is more due to a lack of linearity, rather than true heteroscedasticity. Hence in that case, the outcome is very borderline, almost pointing to quasi-homoscedasticity. The only clear case of heteroscedasticity among the 11 is Winter 2021. Therefore, in the end, one must be very cautious about doubting that the residuals considered constitute a clear situation of heteroscedasticity; rather, the opposite is true.

Now let us move on to autocorrelation Figures 5-8 display the ACF and PCF correlograms used to test for autocorrelation. The confidence bands, calculated using the Bartlett formula, are at a 95% confidence level. Significant autocorrelation is indicated when multiple points fall outside these bands. Our results show that in all diagrams, the points remain within the confidence bands, suggesting a general absence of autocorrelation. The few exceptions are the first points of each seasonal segment. This is not a sign of a larger issue; it is simply because these initial points serve as the starting point for subsequent trends and are therefore inherently correlated with the following data.

**Figure 5.**
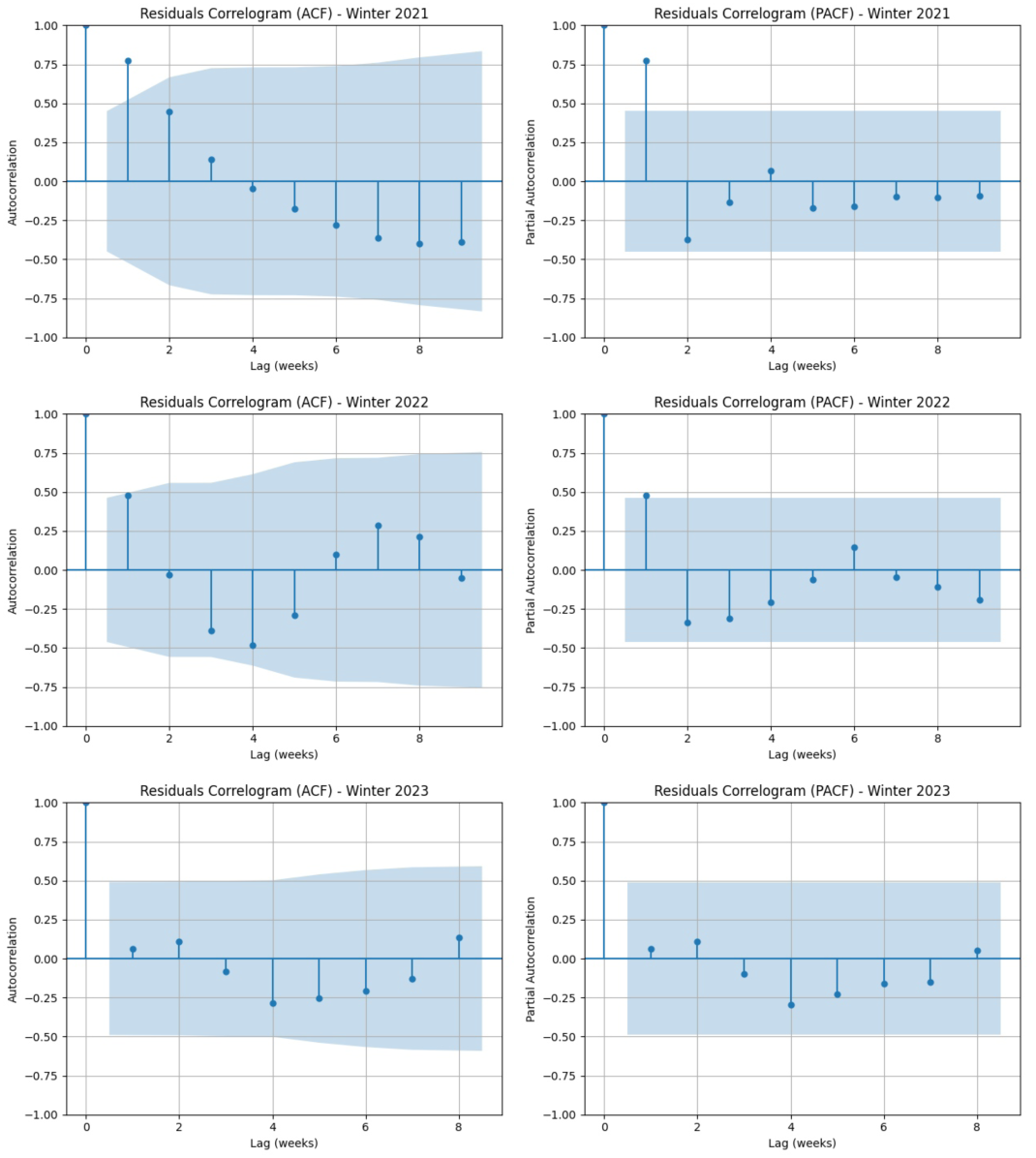
Correlograms for autocorrelation: Winter 2021 (Top-left ACF – Top-right PCF), Winter 2022 (Middle-left ACF – Middle-right PCF), Winter 2023 (Bottom-left ACF – Bottom-right PCF).

**Figure 6.**
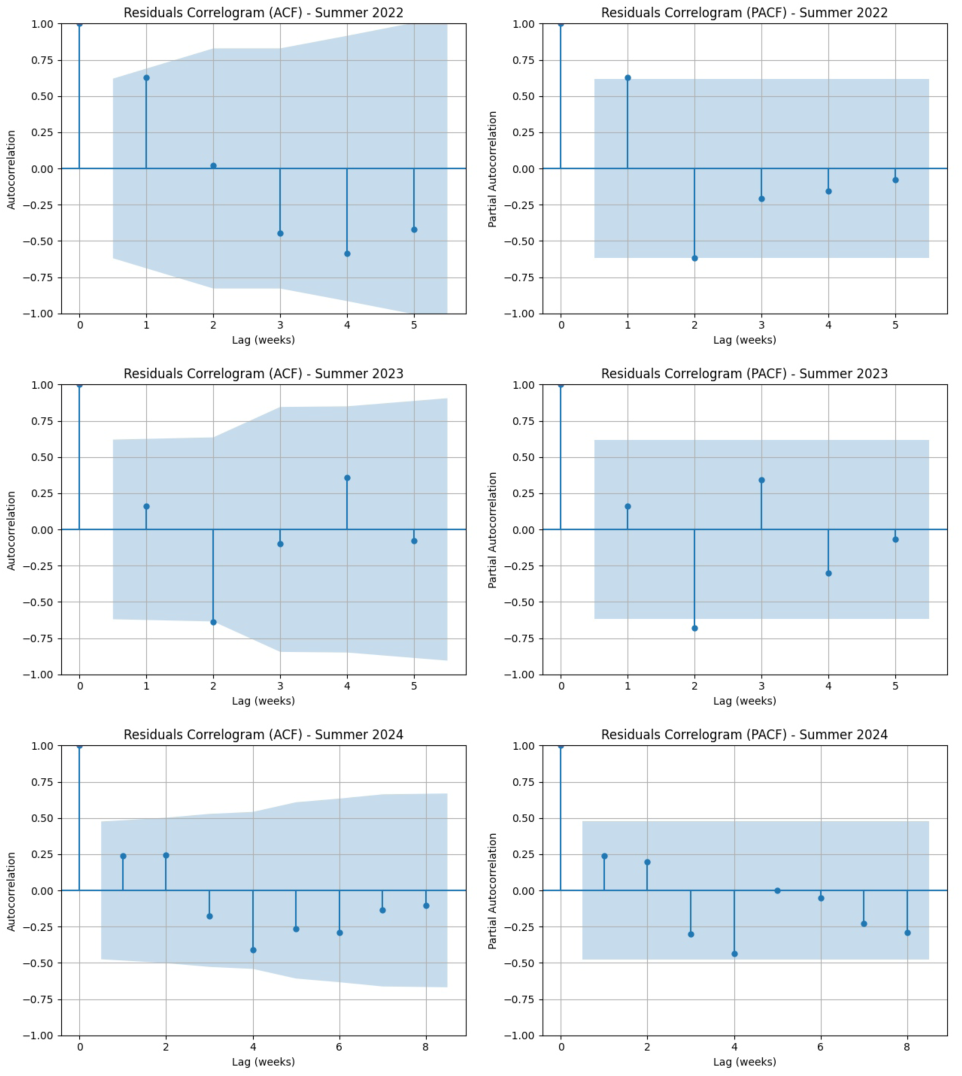
Correlograms for autocorrelation: Summer 2022 (Top-left ACF – Top-right PCF), Summer 2023 (Middle-left ACF – Middle-right PCF), Summer 2024 (Bottom-left ACF – Bottom-right PCF).

**Figure 7.**
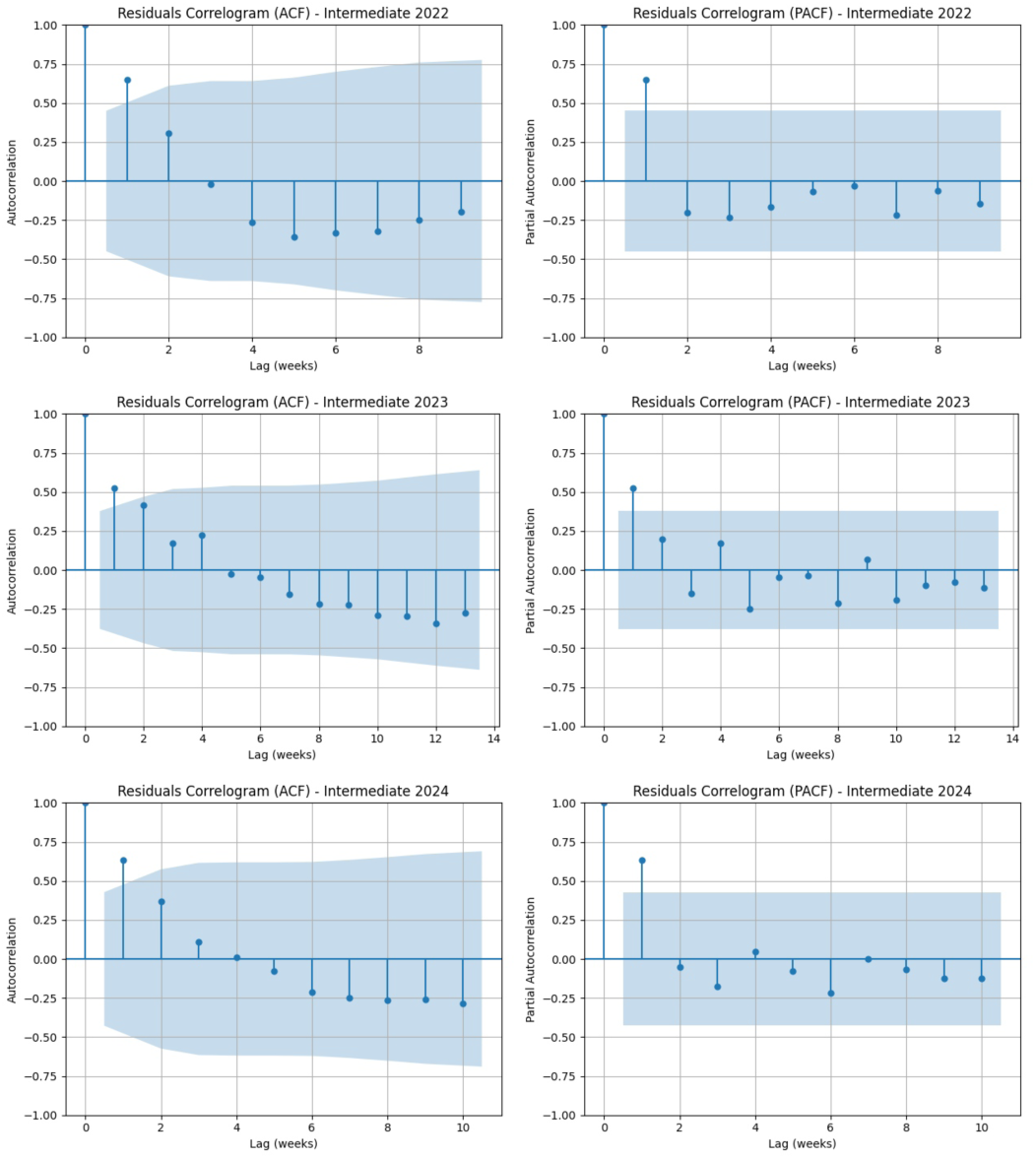
Correlograms for autocorrelation: Intermediate 2022 (Top-left ACF – Top-right PCF), Intermediate 2023 (Middle-left ACF – Middle-right PCF), Intermediate 2024 (Bottom-left ACF – Bottom-right PCF).

**Figure 8.**
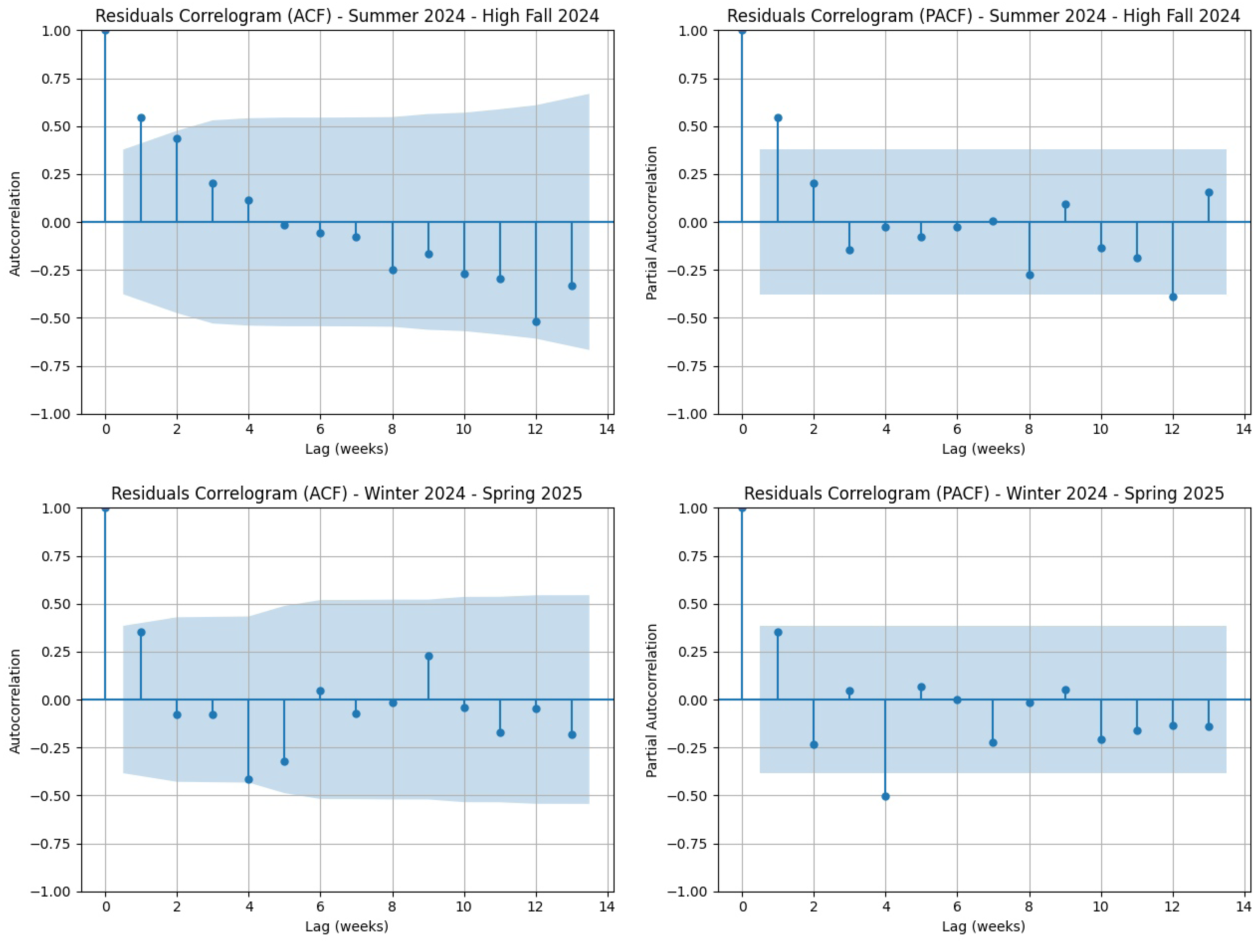
Correlograms for autocorrelation: Summer-Winter 2024-25 (Top-left ACF – Top-right PCF), Extended Spring 2025 (Bottom-left ACF – Bottom-right PCF).

We believe it is helpful to the reader to conclude this Section on the results of statistical tests for the correct applicability of a linear regression model for COVID-19 mortality (2021-2025) by providing a summary Table 9 with the outcomes of the tests for all 11 periods studied with respect to the three dimensions of interest (normality, homoscedasticity and absence of autocorrelation).

**Table 9.** Summary of testing results for normality, homoscedasticity and absence of autocorrelation of linear regression residuals relative to the 11 seasonal segments of the period 2021 – 2025.

| Segment | Normality | Homoscedasticity | No correlation |
| --- | --- | --- | --- |
| Winter 2021 | **Nearly** | Yes | Yes |
| Intermediate 2022 | Yes | **No** | Yes |
| Summer 2022 | Yes | Yes | Yes |
| Winter 2022 | Yes | Yes | Yes |
| Intermediate 2023 | **No** | Yes | Yes |
| Summer 2023 | Yes | Yes | Yes |
| Winter 2023 | **Nearly** | Yes | Yes |
| Intermediate 2024 | **Nearly** | **Nearly** | Yes |
| Summer 2024 | Yes | Yes | Yes |
| Summer-Winter 2024/25 | Yes | Yes | Yes |
| Extended Spring 2025 | Yes | Yes | Yes |

### 3.2. Results from the GLM Poisson Model for COVID-19 Mortality Data in Italy (2021-2025)

We here provide the results of the Poisson GLM model fit to the weekly COVID-19 mortality data for all 11 seasonal segments identified in Section 2. The results are presented in the form of four Figures (9-12) where: Figure 9 shows the Poisson regression curves for all winter seasonal segments, Figure 10 for all summer seasonal segments, Figure 11 for all intermediate seasonal segments, and finally Figure 12 for the two seasonal segments of the period from May 2024 to May 2025.

**Figure 9.**
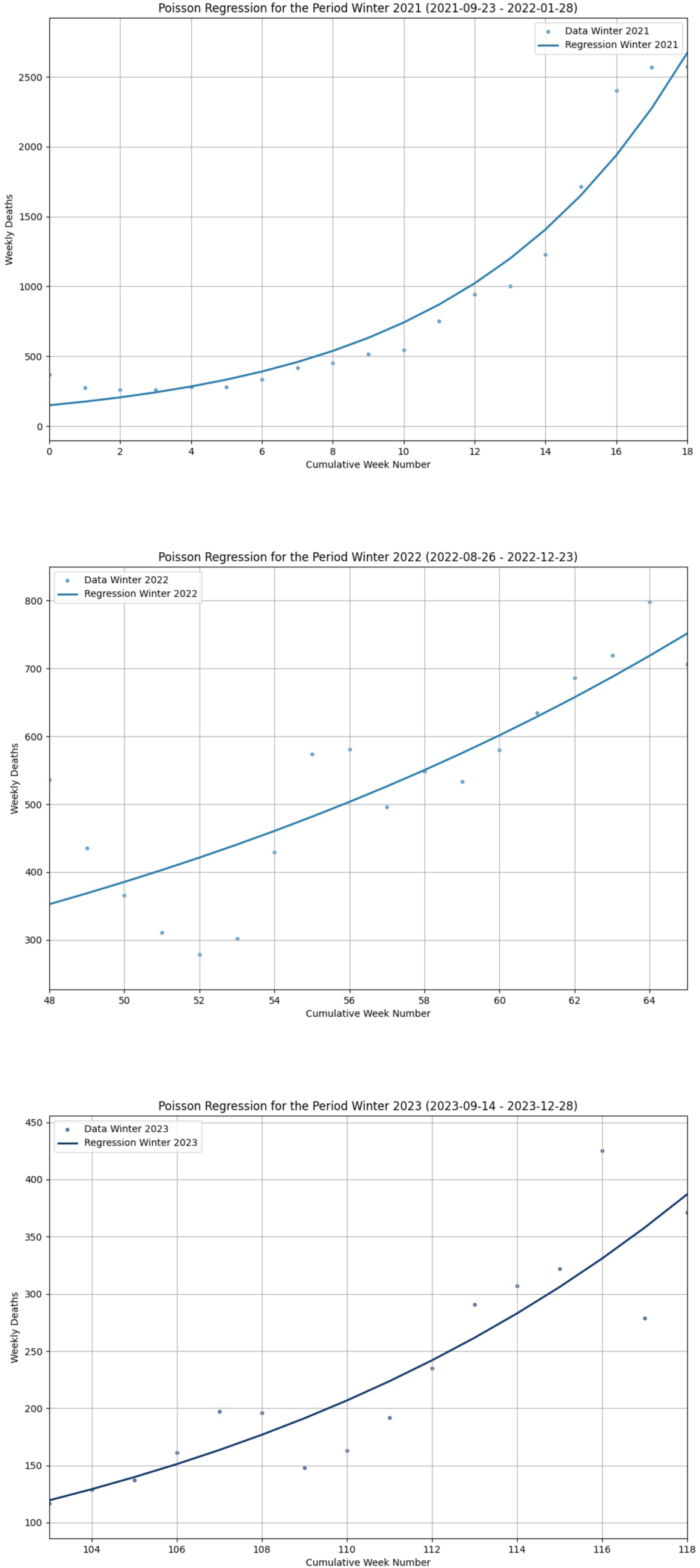
GLM Poisson regression curves for Winter segments. Top: Winter 2021, (*Ŷ* × (exp(*β_1_*) – 1)) = 157.30, Pseudo R^2^ = 0.93. Middle: Winter 2022, (*Ŷ* × (exp(*β_1_*) – 1)) = 23.79, Pseudo R^2^ = 0.55. Bottom: Winter 2023, (*Ŷ* × (exp(*β_1_*) – 1)) = 18.81, Pseudo R^2^ = 0.70.

**Figure 10.**
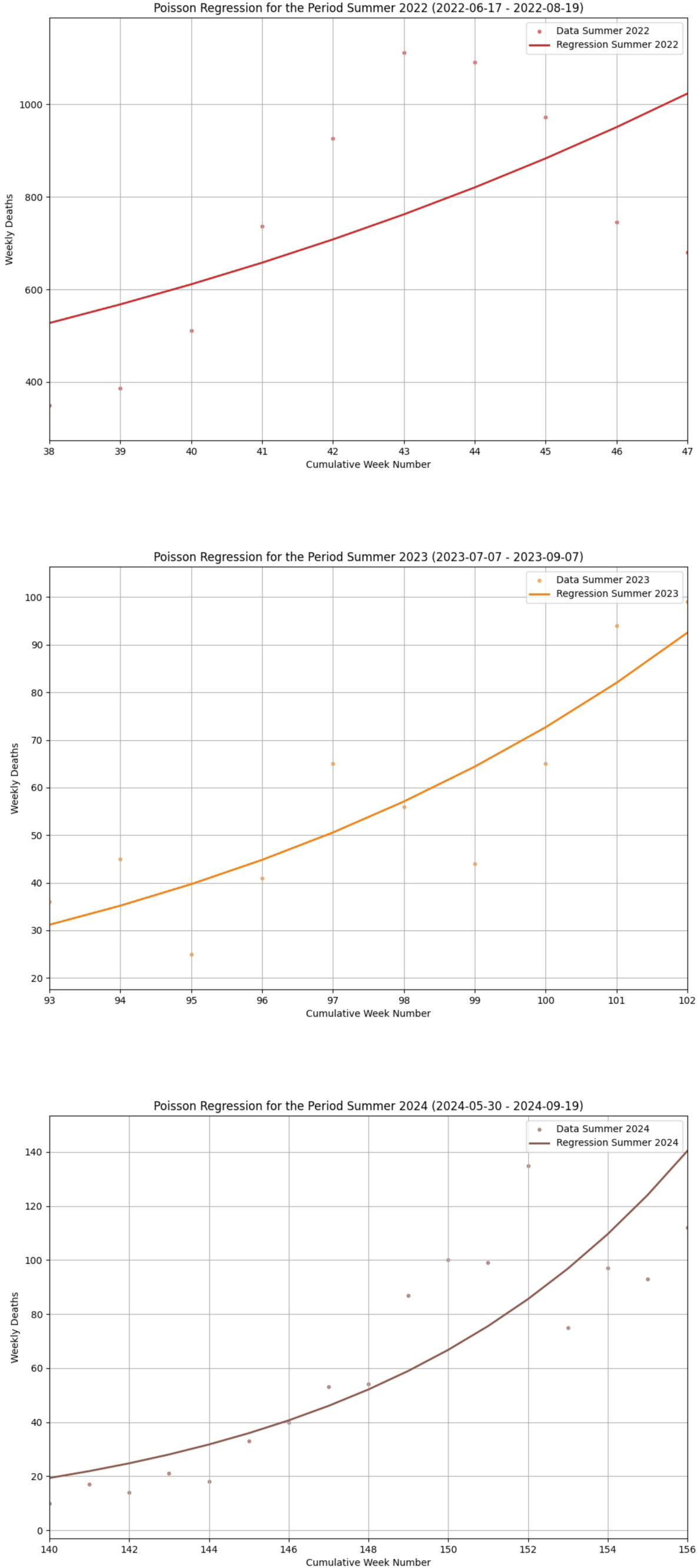
GLM Poisson regression curves for Summer segments. Top: Summer 2022, (*Ŷ* × (exp(*β_1_*) – 1)) = 57.38, Pseudo R^2^ = 0.32. Middle: Summer 2023, (*Ŷ* × (exp(*β_1_*) – 1)) = 7.33, Pseudo R^2^ = 0.45. Bottom: Summer 2024, (*Ŷ* × (exp(*β_1_*) – 1)) = 8.21, Pseudo R^2^ = 064.

**Figure 11.**
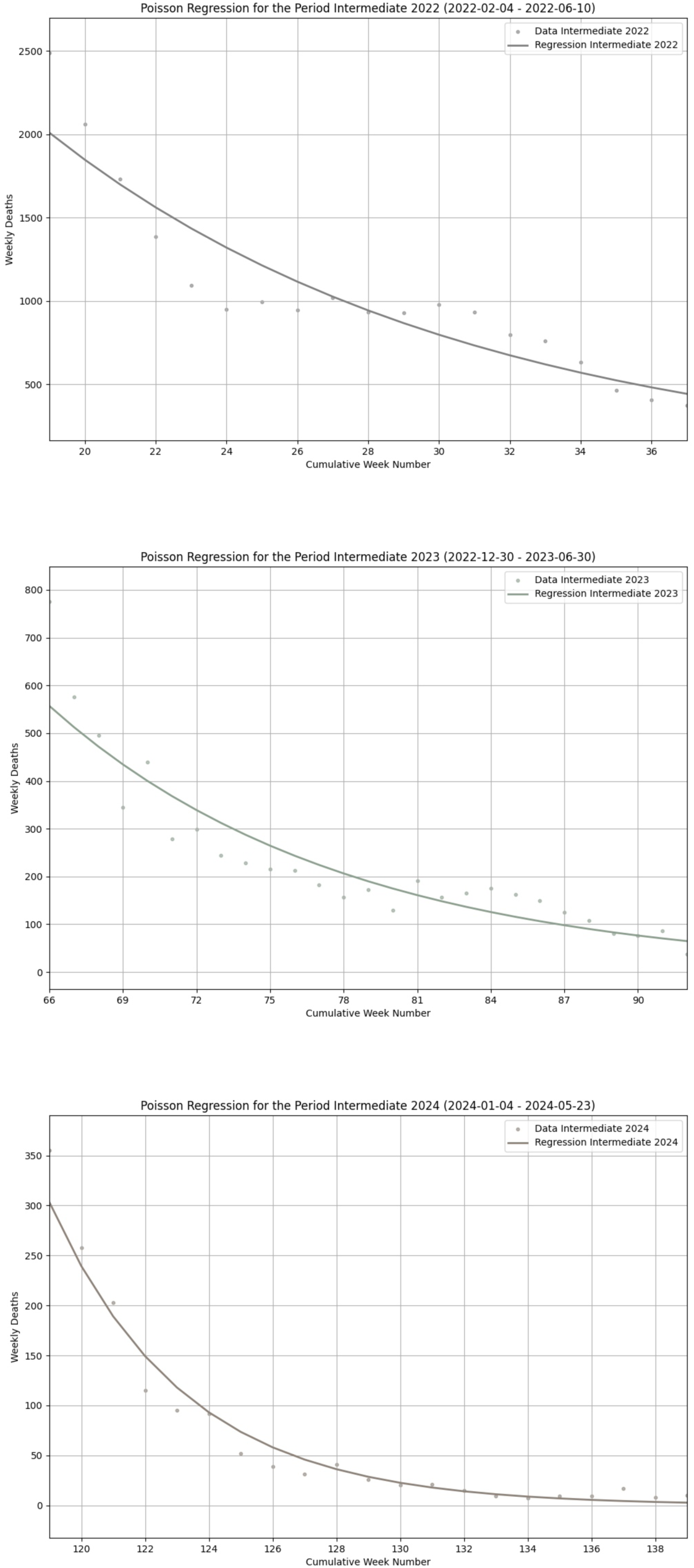
GLM Poisson regression curves for Intermediate segments. Top: Intermediate 2022, (*Ŷ* × (exp(*β_1_*) – 1)) = -84.23, Pseudo R^2^ = 0.84. Middle: Intermediate 2023, (*Ŷ* × (exp(*β_1_*) – 1)) = -18.42, Pseudo R^2^ = 0.82. Bottom: Intermediate 2024, (*Ŷ* × (exp(*β_1_*) – 1)) = -14.35, Pseudo R^2^ = 0.90.

**Figure 12.**
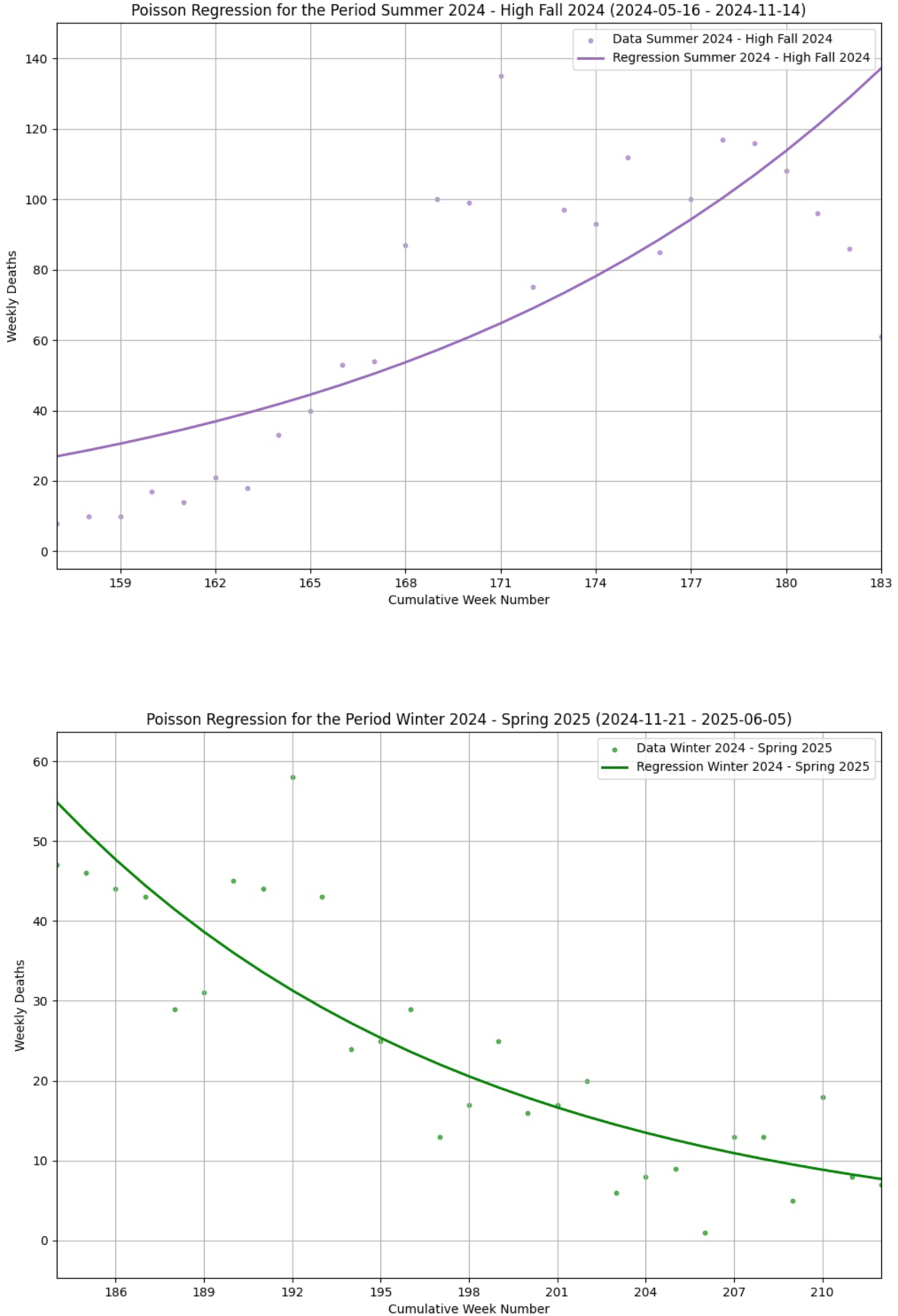
GLM Poisson regression curves for the period May 2024 – May 2025. Left: Summer-Winter 2024/25, (*Ŷ* × (exp(*β_1_*) – 1)) = 4.41, Pseudo R^2^ = 0.49. Right: Extended Spring 2025, (*Ŷ* × (exp(*β_1_*) – 1)) = -1.85, Pseudo R^2^ = 0.45.

Each Poisson regression curve in the Figures is accompanied by its corresponding (*Ŷ* × (exp(*β_1_*) – 1)) value which, starting from the *β_1_* value of the Poisson regression, and applying the transformation with the link function and the average number of deaths for that segment, yields a parameter comparable to the *β_1_* produced by the linear regression model. Also included in the caption of each Figure is the Pseudo R^2^ value which represents the goodness of fit of the model.

After showing the Poisson regression curves in the aforementioned Figures, we present in Table 10 the corresponding values of the weekly COVID-19 death forecasts calculated according to the Poisson model. We defer to the Discussion Section a precise comparison between the results of the linear regression and Poisson models. This comprehensive analysis will be based on: a) a comparison of the growth/decline trends of the profiles identified by lines in one model and curves in the other, b) a comparison based on their respective *β_1_* values, and finally, a numerical comparison of the death forecasts from the two models versus the observed deaths.

**Table 10.**
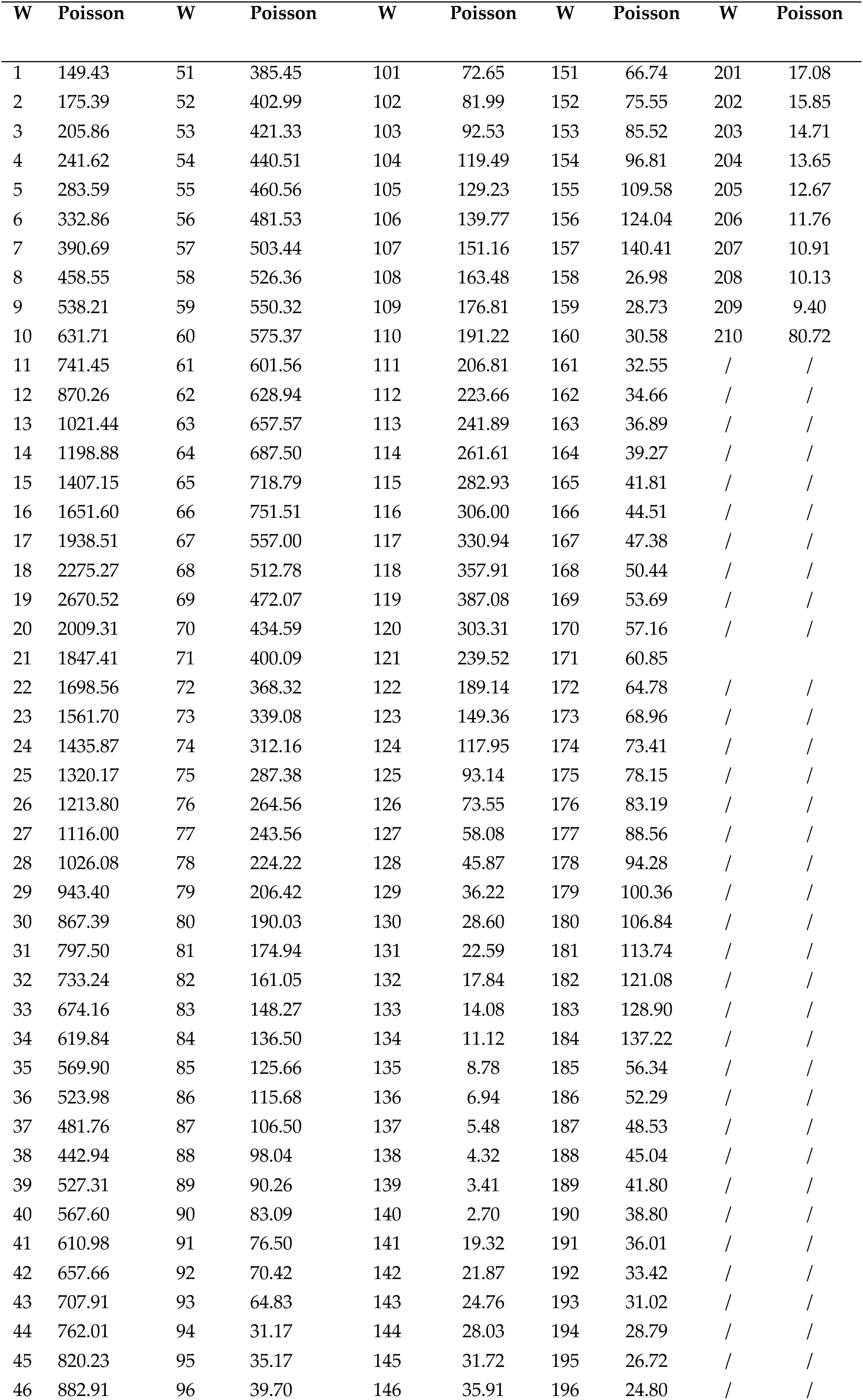
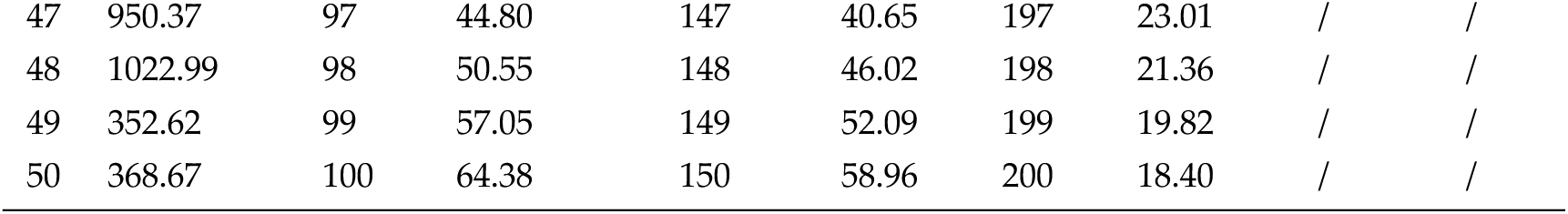
GLM Poisson regression. Predicted weekly deaths from COVID-19: 2021-2025 (210 weeks/W).

Before providing, after the 11 Poisson regression curves, the predicted values of weekly COVID-19 deaths from the Poisson model for all the 11 seasonal segments above, it is impossible not to anticipate, at this point in the treatment, that even with the Poisson model, all three winter curves show an increasing mortality trend, as do the summer curves, albeit less pronounced, while the intermediate periods show a decreasing trend. This is exactly the same type of result provided by the linear regression model in [8]. Similarly, the growth and decline segments observed in the two periods that make up the timeframe studied in [9] from May 2024 to May 2025, show the same trends as the Poisson model curves in Figure 12. Identical statements could be made about the *β_1_* values of the linear model and the linearized Poisson ones, which show a high degree of coincidence.

Now, however, we provide in the Table 10 all the COVID-19 death predictions from the Poisson model for each of the 210 weeks studied.

To close this section, which has first shown the results of the statistical validation tests for the applicability of the linear regression model to COVID-19 mortality data in the extensive time segment from September 2021 to May 2025, and then the results of a similar development exercise on the same mortality data for a Poisson GLM model, we reiterate that the first set of results (statistical tests) has substantially confirmed the possibility of applying a linear model to that data without excessive errors. Meanwhile, the results from the Poisson model show a remarkable alignment with the analogous results of the linear model presented in [8, 9] from every conceivable angle: in the slopes of the curves for all seasonal segments, in the corresponding *β_1_* parameters and in the over 200 forecasts.

However, we believe it is useful to dedicate an entire Section, the next one, to discussing this comparison, performing it with the precision and accuracy that the topic deserves.

## 4. Discussion

The true core of this present work was to find a confirmation, peacefully accepted by experts, for the somewhat surprising results published in [8, 9]. Those results showed that in Italy, during the period 2021-2025, the seasonal profiles of COVID-19 mortality had a growing trend in both winter and summer, with greater emphasis on winter, and then descending profiles in the intermediate period that follows the most central part of winter until late spring. The problem, however, was that those results were obtained by fitting a linear regression model, while it is undeniable that death count problems should be modeled with Poisson or Negative Binomial GLMs. Although there was a rationale behind the use of the linear model, namely, to express the growth and decline trends in a visually clear and unmistakable way, as only straight lines and their slopes can do, those results needed a confirmation that could be fully convincing.

This confirmation has been partially obtained with the results of the statistical tests presented in first part of the previous Section 2, which confirmed that a linear regression model could be applied to that particular time series without committing excessive errors. However, this was not enough, and we developed a more canonical and commonly accepted Poisson GLM model on the same mortality data. The results shown in the second part of Section 2 were confirmatory and surprising afresh.

To discuss them better, we first present the summary Table 11. It takes the 210 observations of COVID-19 deaths in Italy (see columns termed Weekly Deaths of Tables 2-5), grouped into the 11 seasonal segments subject to the model fitting, and numerically compares them with: i) the predictions of the linear model (see columns termed Prediction of Tables 2-5) and ii) with the predictions of the Poisson model (shown in Table 10), calculating the relative MAE and percentage error, for each comparison.

**Table 11.** Comparison between COVID-19 deaths and predictions from linear regression and Poisson regression, with corresponding MAE and percentage errors.

| Seasonal Segment | Deaths | MAE (Linear) | MAE (Poisson) | Percentage Error (Linear) | Percentage Error (Poisson) |
| --- | --- | --- | --- | --- | --- |
| Winter21 | 17183 | 337.47 | 128.45 | 1.96% | 0.75% |
| Intermediate22 | 19883 | 190.36 | 157.55 | 0.96% | 0.79% |
| Summer22 | 7510 | 189.00 | 201.25 | 2.52% | 2.68% |
| Winter22 | 9515 | 66.16 | 62.77 | 0.69% | 0.66% |
| Intermediate23 | 6264 | 67.81 | 45.23 | 1.08% | 0.72% |
| Summer23 | 570 | 11.47 | 9.52 | 2.01% | 1.67% |
| Winter23 | 3670 | 30.47 | 28.26 | 0.83% | 0.77% |
| Intermediate24 | 1432 | 45.53 | 11.72 | 3.18% | 0.82% |
| Summer24 | 1058 | 13.01 | 16.84 | 1.23% | 1.59% |
| Wint-Sum24-25 | 1845 | 17.40 | 22.11 | 0.94% | 1.20% |
| Ext-Spring25 | 671 | 5.91 | 6.65 | 0.88% | 0.99% |
| Total<br>(Average per period) | 69601<br>(6327) | **88.60** | **62.76** | **1.48%** | **1.15%** |

Table 11 clearly shows that the MAE of the Poisson regression is only slightly better than that of the linear regression, 62.76 versus 88.60 (the lower the MAE, the more accurate the prediction). If we then look at the percentage errors, we see that the quantity of errors on this huge number of predictions (averaged over the number of deaths per period) is indeed lower with the Poisson model, at exactly 1.15%. However, moving to the linear regression model results in a shift to 1.48%, which is frankly negligible. It is still important to remember, though, that out of the 210 predictions, the linear regression model provided 7 negative values, which, although very few, are obviously forbidden as they are meaningless.

An even more interesting comparison is to look at the slopes of the Poisson curves versus the lines of the linear regression model. We have already documented that the trends match 100% across all 11 periods, that is when the linear model shows an ascending trend, so does the Poisson model, and similarly for descending trends. However, a more compelling comparison is between the slopes of the Poisson curves and the lines for each seasonal segment.

Table 13 shows the *β_1_* values of the linear model (second column) for each period and the corresponding linearized *β_1_* values of the Poisson model (fourth column). It is possible to see that the two series of values are practically identical, with such minimal differences that they do not even warrant being counted.

**Table 12.** Comparison between the slopes of the linear model and the Poisson model, with the linearized Poisson *β_1_*.

| Seasonal Segment | Linear $\beta_l$ (slope) | Poisson $\beta_l$ | Poisson $(\hat{Y} \cdot (\exp(\beta_l) - 1))$ (slope) |
| --- | --- | --- | --- |
| Winter21 | 126.45 | 0.1602 | 157.3 |
| Intermediate22 | -84.38 | -0.0840 | -84.23 |
| Summer22 | 54.80 | 0.0736 | 57.38 |
| Winter22 | 23.28 | 0.0445 | 23.79 |
| Intermediate23 | -17.76 | -0.0827 | -18.42 |
| Summer23 | 6.72 | 0.1209 | 7.33 |
| Winter23 | 17.52 | 0.0784 | 18.81 |
| Intermediate24 | -11.89 | -0.2361 | -14.35 |
| Summer24 | 7.20 | 0.1240 | 8.21 |
| WinterSummer2425 | 4.08 | 0.0625 | 4.41 |
| ExtendedSpring25 | -1.81 | -0.0746 | -1.85 |

Ultimately, this is the truly surprising and, we believe, innovative aspect of this study: it has shown that, at least in the limited, yet temporally extended, circumstances analyzed, a simpler and easier-to-understand linear model provides the same results as a Poisson model, especially concerning the seasonal growth and decline trends of the COVID-19 mortality subject of our interest.

To conclude, we present Figure 13 which displays the COVID-19 mortality data from 2021 to 2025 with superimposed curves from the Poisson model for each seasonal segment (each distinguished by a different color), along with an inset box showing the linear model lines from study [8]. This last Figure perfectly exemplifies the overlap of the two models’ results, effectively summarizing the most significant data that has emerged from the current study.

**Figure 13.**
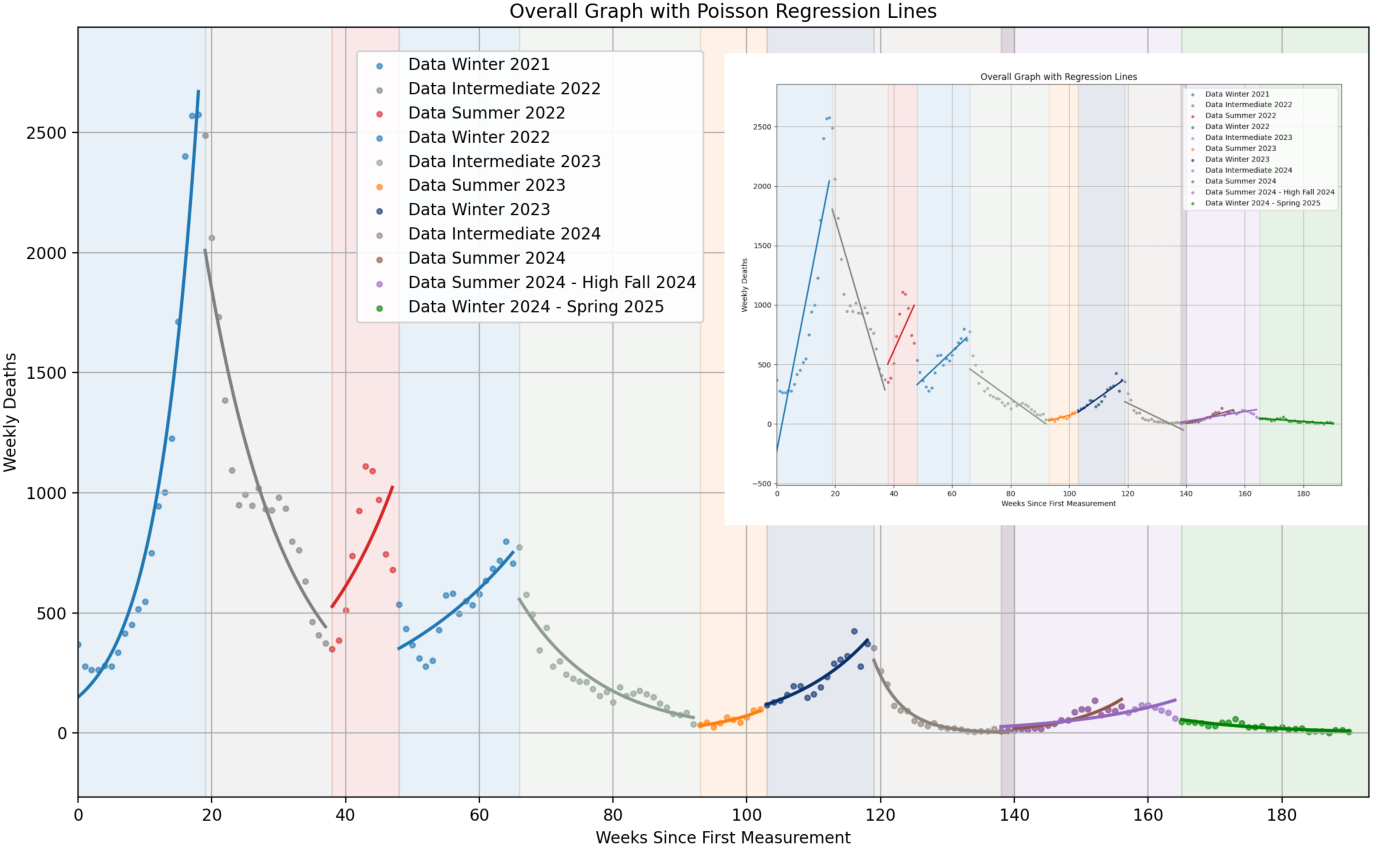
In the big picture, the COVID-19 death data for the entire 2021-2025 period with the Poisson model curves superimposed, with each distinct seasonal segment represented by a different color. In the small inset, an analogous plot with the linear model lines, inspired by [8].

Of course, we do not forget the limitations of the current study. The first of these, we must remember, is the clear preference for regression models based on Poisson or negative binomial distributions when dealing with count data, given that the case examined here is only a single event, however significant it has been.

We also cannot avoid concisely listing a summary of the limitations that concern not so much the modeling discussions conducted so far but those related to the content, namely the observation of COVID-19 mortality during the period of interest.

First, this study’s findings are limited to the specific epidemiological context of Italy between September 2021 and May 2025, a period dominated by Omicron and post-Omicron SARS-CoV-2 variants. The results might differ in other periods or geographic locations with different circulating lineages [20].

Another key limitation with this kind of studies, already discussed at length in [8], is they deliberately avoid identifying the causes behind the observed seasonal mortality trends. While these trends could be influenced by various factors like climate, social behaviors, immunity levels, and public health measures, the decision was made to focus purely on observing and quantifying the trends themselves [21-26]. This observational approach was chosen due to the unreliability of relevant data during this period, such as vaccination rates and recorded infections. Discussing the causes without reliable data would have been just speculative.

Finally, the use of Italian government data presents its own set of limitations. The data, provided as aggregated measures from two different sources, has been subject to corrections and adjustments over time, only recently achieving relative stability. Nevertheless, public thanks must be given to the Italian institutions, because they have still guaranteed, and continue to do so, a constant flow of data that allows independent researchers to continue studying the phenomenon of COVID spread, even in the current times.

## 5. Conclusions

In the field of epidemiological modeling, Poisson GLM models are considered the standard for analyzing count data, such as deaths, and are extensively used in epidemiology and beyond [27-36].

However, our research [8, 9] explored the use of a simple linear regression on weekly COVID-19 mortality data in Italy (2021-2025). The results revealed a significant finding: despite a general downward trend, we consistently observed an increase in mortality during both winter and summer. The question we asked in the present paper was whether linear regression could still be a valid tool for highlighting the slopes of these seasonal trends. Our direct comparison with a Poisson GLM model provided convincing answers.

The key findings of this current analysis, in fact, have been: As to statistical validity, hypothesis tests confirmed that linear regression was applicable to this specific time series, as its conditions for normality of residuals, homoscedasticity, and lack of autocorrelation were substantially met. As to trend correspondence, there was a 100% alignment between the two models in identifying growth or decline trends across all 11 seasonal periods. Finally, as to accuracy, both models showed exceptional precision in forecasting deaths. The Poisson model had a slightly lower MAE (62.76) compared to the linear one (88.60). In percentage terms, the Poisson model’s error was 1.15%, while the linear regression’s error was just 1.48%, a negligible difference.

In conclusion, our current study has shown that in this specific context, a simpler and more visually intuitive linear regression model provided the same results and nearly identical accuracy as the more complex Poisson model. This suggests that, under certain circumstances, linear regression can be an effective tool for understanding mortality trends.

## Supplementary Materials

No further supporting information.

## Author Contributions

Conceptualization, M.R.; methodology, M.R.; software, G.D.; validation, M.R. and G.C.; formal analysis, G.C.; investigation, M.R.; resources, M.R.; data curation, M.R. and G.C.; writing—original draft preparation, M.R.; writing—review and editing, M.R. and E.D.; visualization, G.C.; supervision, M.R.; project administration, M.R.. All authors have read and agreed to the published version of the manuscript. Both authors have contributed substantially to the work reported.

## Institutional Review Board Statement

This study uses publicly available, aggregated data that contains no private information. Therefore, ethical approval is not required.

## Informed Consent Statement

Not applicable: Neither humans nor animals nor personal data are involved in this study.

## Data Availability Statement

All the initial COVID-19 deaths data are downloadable from two public, open access repositories, specifically: i) the repository maintained by the Italian Civil Protection Department, under the Italian Presidency of the Council of Ministers (https://github.com/pcm-dpc/COVID-19/blob/master/dati-andamento-nazionale), and ii) the repository maintained by the Italian Ministry of Health, (https://www.salute.gov.it/new/it/tema/covid-19/report-settimanali-covid-19/). Moreover, all the data on which both the statistical hypothesis tests and the Poisson regression model are based have been made fully available in Section 2 under a tabular format. All the results of this study are fully reproducible by using the methods described in this paper and the data made available above. Further reasonable requests relative to data and code can be also addressed to the corresponding author.

